# Assessing the Drivers of Wasting among Children Under 5 and Their Mothers in The Bay and Hiran Regions of Somalia

**DOI:** 10.1101/2025.02.21.25322675

**Authors:** Samantha Grounds, Shelley Walton, Kemish Kenneth Alier, Sydney Garretson, Said Aden Mohamoud, Sadiq Abdikadir, Qundeel Khattak, Mohamud Ali Nur, Abdullahi Muse Mohamoud, Meftuh Omer, Mohamed Billow Mahat, Adam Abdulkadir Mohamed, Abdifatah Mohamed, Marina Tripaldi, Nadia Akseer

**Affiliations:** Johns Hopkins University.; Save the Children Somalia; Save the Children International; Somalia Ministry of Health

**Author notes:** Corresponding Author: Nadia Akseer Associate Scientist, Department of International Health Johns Hopkins University.

**Keywords:** wasting, acute malnutrition, children under 5, humanitarian

## Abstract

**Background:** To address Somalia’s high burden of wasting, it is imperative to understand the country’s context-specific drivers of wasting. This study assessed the drivers of wasting among children under 5 (CU5) and mothers in Somalia’s Bay and Hiran regions to inform strategies to address wasting.

**Methods:** The data comes from the midline (September 2023) and endline (December 2023) data collection of a randomized controlled trial. Child and maternal outcomes (weight-for-height z-scores (WHZ) and mid-upper arm circumferences (MUAC), respectively) were explored continuously for children via linear regression and as binary outcomes via Poisson regression for children and mothers. A hierarchical model building approach was used, mapping variables into the basic, intervention, underlying, and immediate levels. Separate midline and endline models were analyzed cross-sectionally, comparing drivers by seasonality, and CU5 models were further stratified by region and age.

**Results:** The burden of CU5 wasting was 12.9% at midline and 14.4% at endline. The following variables were drivers of low WHZ across different models: child illness, open defecation, low maternal MUAC, no maternal education, having a male-headed household, and living in a household without joint decision-making. Egg/flesh food consumption and higher maternal MUAC were protective of WHZ. Wasting among mothers was 8% at midline and 12% at endline. Household food insecurity, open defecation, and poor waste disposal practices were drivers of mothers’ wasting, whereas maternal decision-making was protective.

**Conclusion:** This study highlights variation in the key drivers of wasting by region, season, and child age and contributes to an expanding body of evidence on the multifactorial drivers of wasting, encouraging context-specific approaches that address the immediate, underlying, and basic causes of malnutrition. The findings emphasize the importance of maternal nutrition for child nutrition outcomes and the need for interventions considering household food security, sanitation, and gender dynamics in this humanitarian setting.

**Registration:** The cluster-RCT is registered at ClinicalTrials.gov, ID: NCT06642012.

## Background

The World Health Organization (WHO) estimates that 45 million children worldwide suffered from wasting in 2022, with close to 14 million experiencing severe wasting (see **Text S1** for technical definition of wasting).[1] In 2022, the estimated global prevalence of wasting among children under 5 (CU5) was 6.8%, with an estimated 12.2 million CU5 in Africa affected by wasting, of which nearly three million were severely wasted.[1] Wasting is not only a concern among young children, but also among pregnant and lactating women (PLWs). Among the 12 countries, including Somalia, most severely impacted by the ongoing global food and nutrition crisis, acute malnutrition among PLWs increased by 25% from 2020 to 2022, totaling nearly 7 million acutely malnourished women and adolescent girls in 2022.[2,3]

Somalia’s 2020 Health and Demographic Survey estimate that 11.6% of CU5 in Somalia are wasted, with 6% of CU5 being severely wasted, and a recent publication estimated that 1.7 million CU5 in Somalia would suffer from acute malnutrition in 2024.[4–6] Approximately 62.6% of CU5 in Bay and approximately 59.6% of CU5 in Hiran were estimated to be acutely malnourished from January-December 2024.[6] Within these regions, Baidoa and Belet Weyne districts show high overall moderate acute malnutrition and severe acute malnutrition median prevalences of 14.2% and 4.9%, respectively, in Baidoa district and of 14.7% and 3.6%, respectively, in Belet Weyne district, falling into the critical/phase 4 category of The Integrated Food Security Phase Classification’s (IPC) acute malnutrition scale.[7,8]

Despite Somalia’s high wasting burden, there is limited primary evidence around the drivers of wasting in Somalia, and the evidence that does exist is outdated.[9] Much of the peer-reviewed primary research evidence comes from national household survey data from 2007-2010.[10–14] These analyses found the following characteristics to be associated with wasting among children 6-59 months of age in Somalia: sex, age, illness, vaccination, dietary and nutritional factors, maternal age and mid-upper arm circumference (MUAC), household size, number of CU5 in the household, environmental and WASH factors, and other demographic and social factors (livelihood, displacement status, urbanization, and conflict).[10–14] More recently, an analysis of the 2019 Somalia Micronutrient Survey identified low household wealth, recent episodes of diarrhea, and iron deficiency as risk factors for wasting among CU5.[15] No peer-reviewed primary research on the drivers of wasting among PLW or mothers of CU5 in Somalia was identified.

This study seeks to address this evidence gap and to inform strategies to address wasting in Somalia by assessing the drivers of wasting among CU5 and their mothers in the Bay and Hiran regions, selected due to their high acute malnutrition burdens.[8,16] Specifically, this study seeks to answer the following research question: What are the top drivers of wasting among children under five and their mothers in the Bay and Hiran regions of Somalia?

Specific objectives of this analysis include:

1. Exploring the drivers of wasting among children under 5 in the Bay and Hiran regions of Somalia, stratifying results by region and by child age (9-23 months versus 24-59 months) and comparing by seasonality.
2. Exploring the drivers of wasting among mothers of children under 5 in the Bay and Hiran regions of Somalia and comparing by seasonality.

## Methods

Data for this study comes from a cluster randomized controlled trial (cRCT) to evaluate the effectiveness of a CashPlus for Nutrition program implemented by Save the Children.[17,18] This study took place in the Bay and Hiran regions in southwest Somalia, a humanitarian setting where flooding, droughts, and conflict have threatened livelihoods, increased food insecurity, and contributed to both displacement from these regions as well as arrivals of internally displaced persons to these regions throughout the years.[19–25] Additional details describing the cRCT and study setting are available in **Text S2** and **S3**.

Baseline (May 2023), midline (September 2023), and endline (December 2023) data were collected during the cRCT.[17,18] Data for this analysis comes from the midline and endline data, corresponding to the transition from the Xagaa dry season to the Deyr rainy season and to the Jilaal dry season, respectively.[26] In October 2023, between the midline and endline data collection points, major flooding occurred in Somalia, with an estimated 2.5 million people affected and thousands displaced.[24,27,28] The baseline data was not included in this analysis, as the main trial was interested in wasting prevention, and, therefore, children were only recruited into the study if they were not malnourished; therefore, a low prevalence of wasting was expected in the baseline data.

The outcome of interest in this analysis is wasting, defined by weight-for-height Z-scores for CU5 and by MUAC for mothers.[29–34] WHZ was calculated using the zscore06 command in STATA, with child wasting defined by a WHZ < -2 standard deviations (SD) from the median age- and sex-specific WHZ from the WHO 2006 Child Growth Module.[29–31,35–37]. Child wasting as a binary outcome was examined at midline and endline with the overall CU5 samples via multivariable Poisson regression models. However, to examine the age- and region-specific drivers of child wasting and due to sample size limitations and the rarity of wasting outcomes when considering these stratified models, WHZ as a continuous outcome was modelled via linear regression models to allow for a broader exploratory analysis of the factors driving child nutritional status. For the mothers’ models, MUAC was categorized as a binary outcome to examine wasting, defined as a MUAC less than 23 centimeters, via multivariable Poisson regression models.[32]

Variables were analyzed cross-sectionally for statistically significant associations with the outcome. While recognizing that the analytical methods used here demonstrate associations and not causality, the term “drivers” is used for simplicity, and, in this paper, drivers are defined as covariates that are correlated with the outcome of interest.

**Table S1** shows the independent variables that were tested. Variables that had complete or less than 15% missing responses and that could plausibly be related to wasting outcomes were considered. Additionally, to address covariate imbalance that may affect regression results, variables that had limited variation in a given sample (using a threshold of more than 75%/less than 25%) were not assessed in the corresponding model. Variables concerning location, wealth and expenditure, displacement, household decision-making and women’s empowerment, intervention components, household food security and environment, health- and nutrition-related knowledge and practices, child diet and disease, and maternal characteristics were considered (see **Table S1** and **S2**). Complete data on household crowding was only available in the midline data, and displacement, head of household, and maternal education data were only collected at endline.

The UNICEF Conceptual Framework on Maternal and Child Nutrition was adapted into a statistical framework for analysis, organizing variables into the basic/enabling, underlying, and immediate levels of determinants of maternal and child nutrition.[38–40] Additionally, intervention-related indicators were mapped into a separate level between the basic/enabling and underlying levels. **Figure 1** shows the framework mapping of variables considered in the CU5 models (see **Figure S1** for mothers).

**Figure 1.**
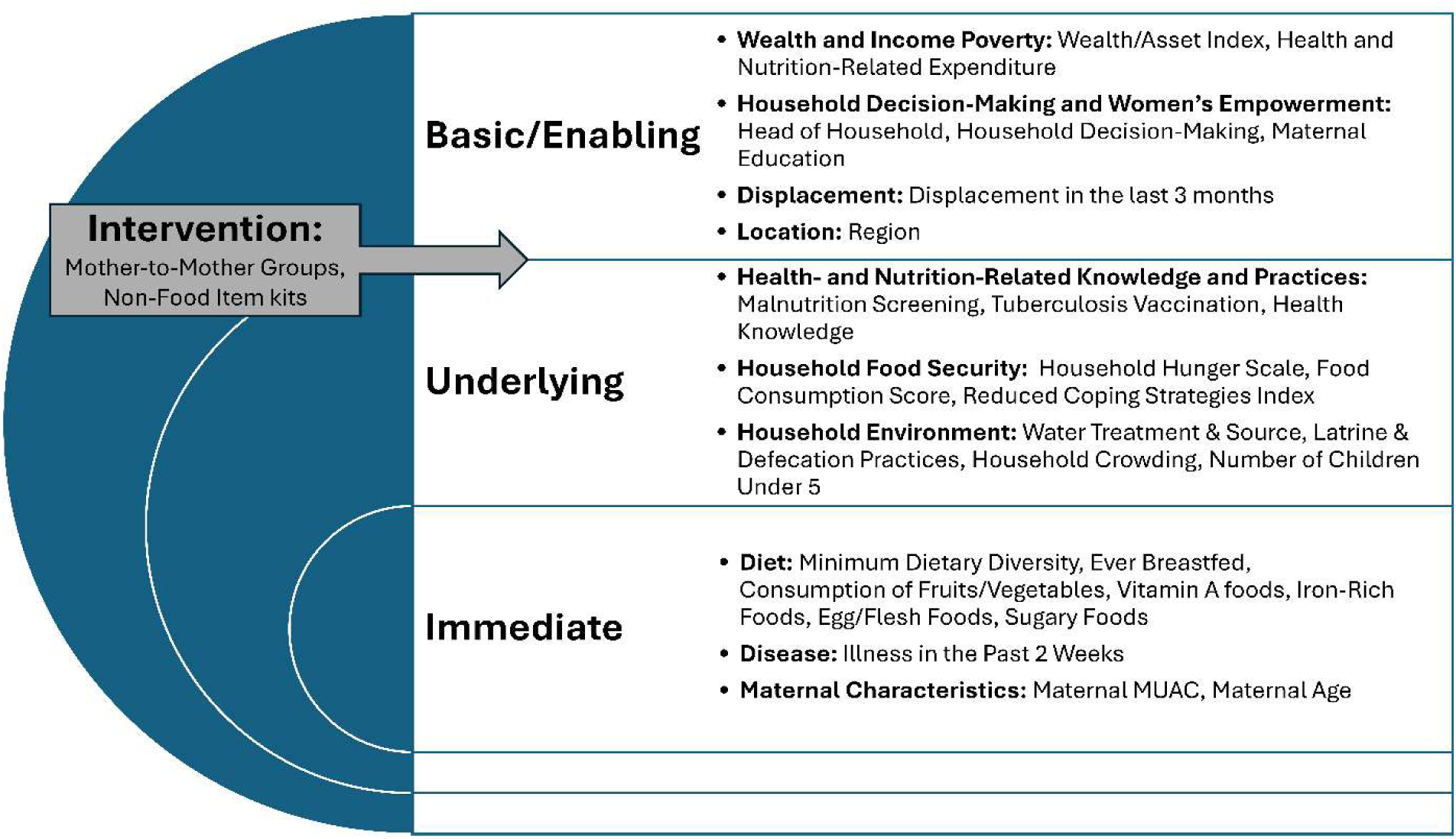
Mapping of Variables Tested in the CU5 Models

This analysis was performed on sub-samples of CU5 and mothers from the cRCT, with 956 children and 1066 mothers at midline and 833 children and 1023 mothers at endline. See **Text S5** for inclusion and exclusion criteria.

The midline and endline data sets were analyzed separately and cross-sectionally, and a hierarchical model-building approach was used.[38–40] During the first stage of analysis, unadjusted bivariate relationships between the outcome and each independent variable were examined. Next, major confounder-adjusted relationships were examined, adjusting for child sex and age, household region, and trial arm in the CU5 models and adjusting for household region and trial arm in the mothers’ models. From these major confounder-adjusted results, any independent variable that had an association with the outcome with a p-value <0.20 was retained for the final stage.[41] A backwards-elimination model building approach was then used and implemented in a hierarchical way at four levels according to the statistical framework.[40] Variables with p-values <0.10 were retained in the final fully adjusted models (see **Text S6** for details). The results presented here focus on the findings from these fully adjusted models, as these are believed to represent the most unbiased and unconfounded results. The cRCT trial arm was adjusted for; however, to not overfit the models, the village cluster was not controlled for in this analysis.

For examining wasting and WHZ among CU5, “overall” models with the entire samples of children were built with both the midline data and endline data (separately). Additional models for WHZ were stratified by region and by child age (9-23 months versus 24-59 months). For mothers’ wasting, only overall models with all mothers were built at midline and endline.

Model fit checks were assessed for all final adjusted models (see **Text S6**). A statistical significance level of α = 0.05 is considered.

Data was analyzed using STATA version 18.5 (StataCorp, College Station, TX, USA).

## Results

### Participant Characteristics

**Table 1** highlights key child, household, and maternal characteristics. Additional characteristics are reported in **Table S3**.

**Table 1.**
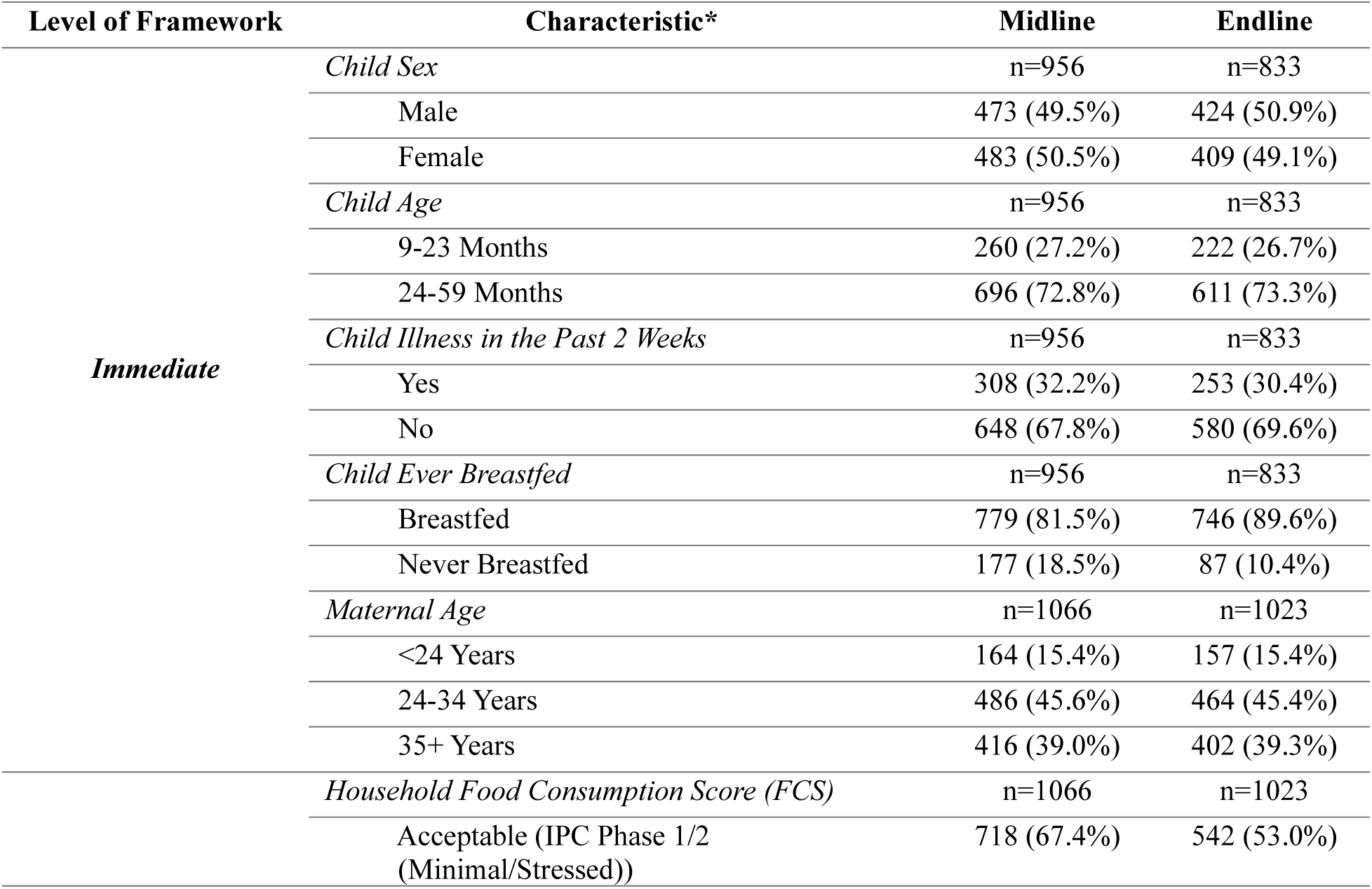
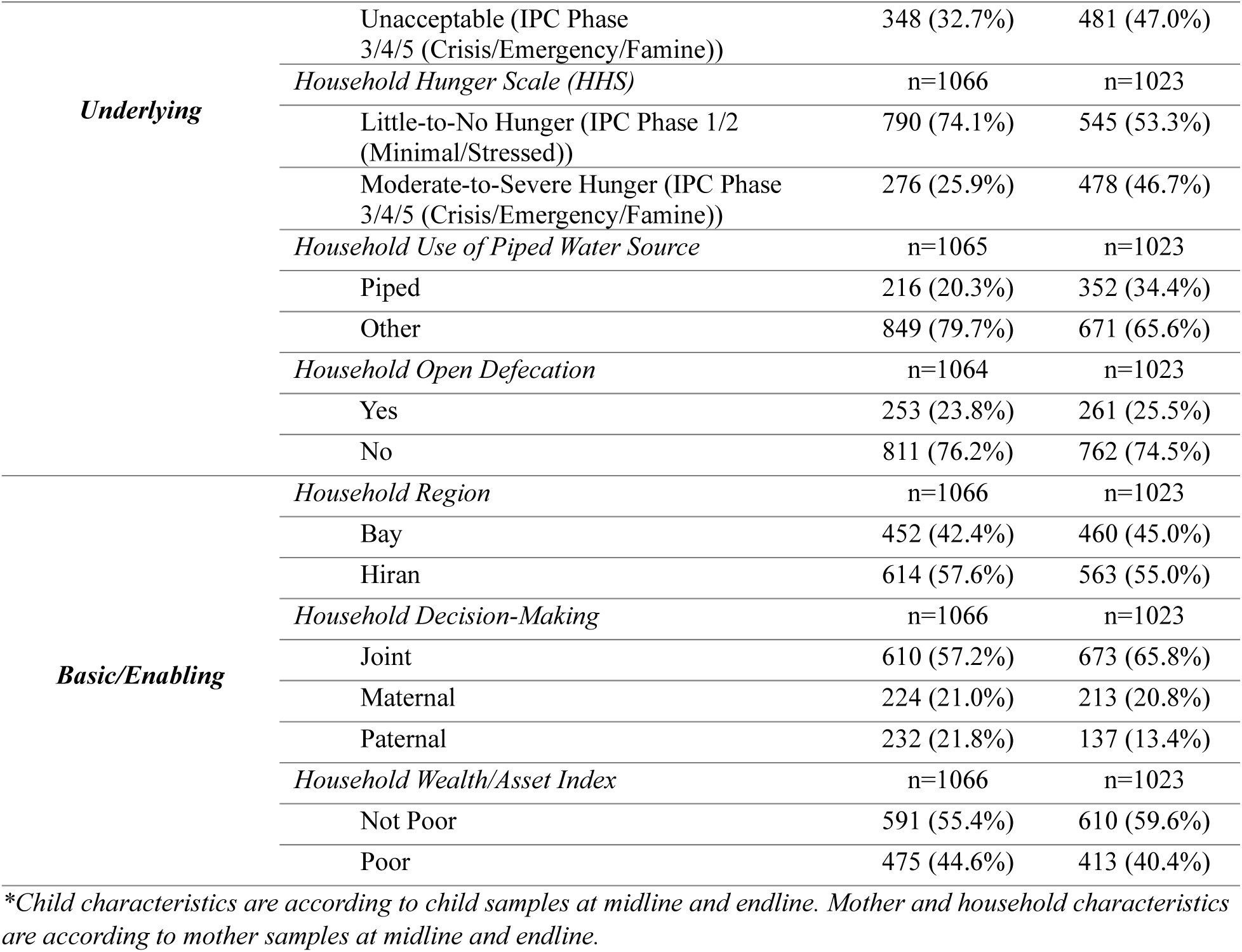
Key Child, Mother, and Household Characteristics*

### Nutrition Status

Child and maternal nutrition status are reported in **Table 2**. Average WHZ was -0.7 SD at midline (95% CI: -0.75, -0.61) and -0.8 SD at endline (95% CI: -0.89, -0.74). Approximately 13% of children were wasted at midline (95% CI: 10.9%, 15.1%) and over 14% wasted at endline (95% CI: 12.2%, 16.9%). At both timepoints, the burden of wasting was higher in Hiran than in Bay and higher among older children than younger children, prompting stratified analyses by region and by age.

**Table 2.**
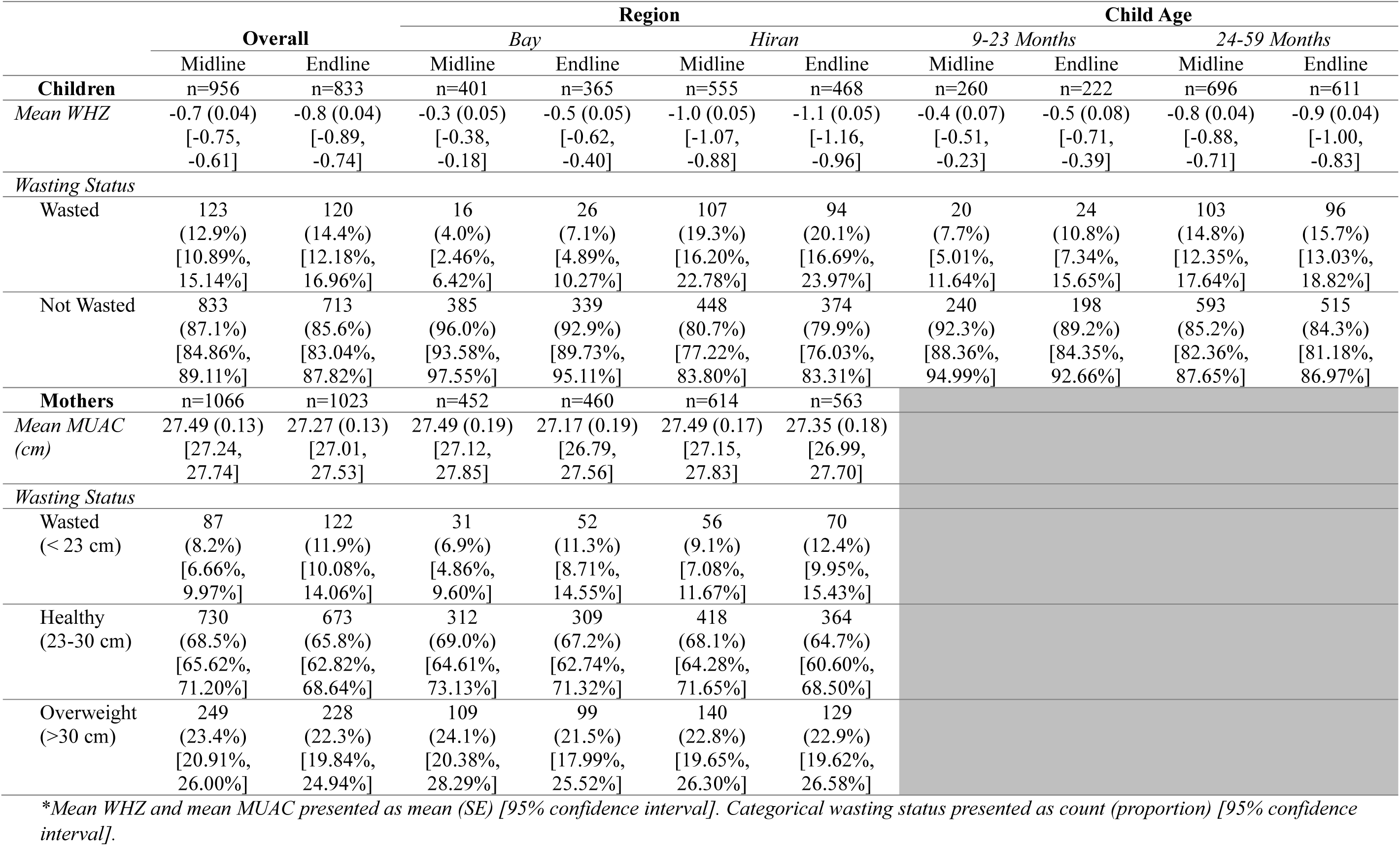
Nutrition Status of Children and Mothers

At midline, around 8% of mothers were wasted (95% CI: 6.7%, 9.9%), whereas approximately 12% were wasted at endline (95% CI: 10.1%, 14.1%). While wasting was slightly higher in Hiran than in Bay among mothers, this difference was not statistically significant (chi-squared p-value = 0.182 at midline and 0.579 at endline).

### Drivers of CU5 Wasting

The presented results are from the final hierarchical multivariable linear regression results for child WHZ; additional indicators were marginally statistically significant or were statistically significant in the crude or basic-adjusted models, and these results are available in the data tables. Results from the multivariable Poisson regression models assessing wasting as a binary outcome are presented in **Tables S4** and **S5**, with relevant findings highlighted here. Figure 2 summarizes the statistically significant drivers of child WHZ from the final hierarchical multivariable regression results across different analyses.

**Figure 2.**
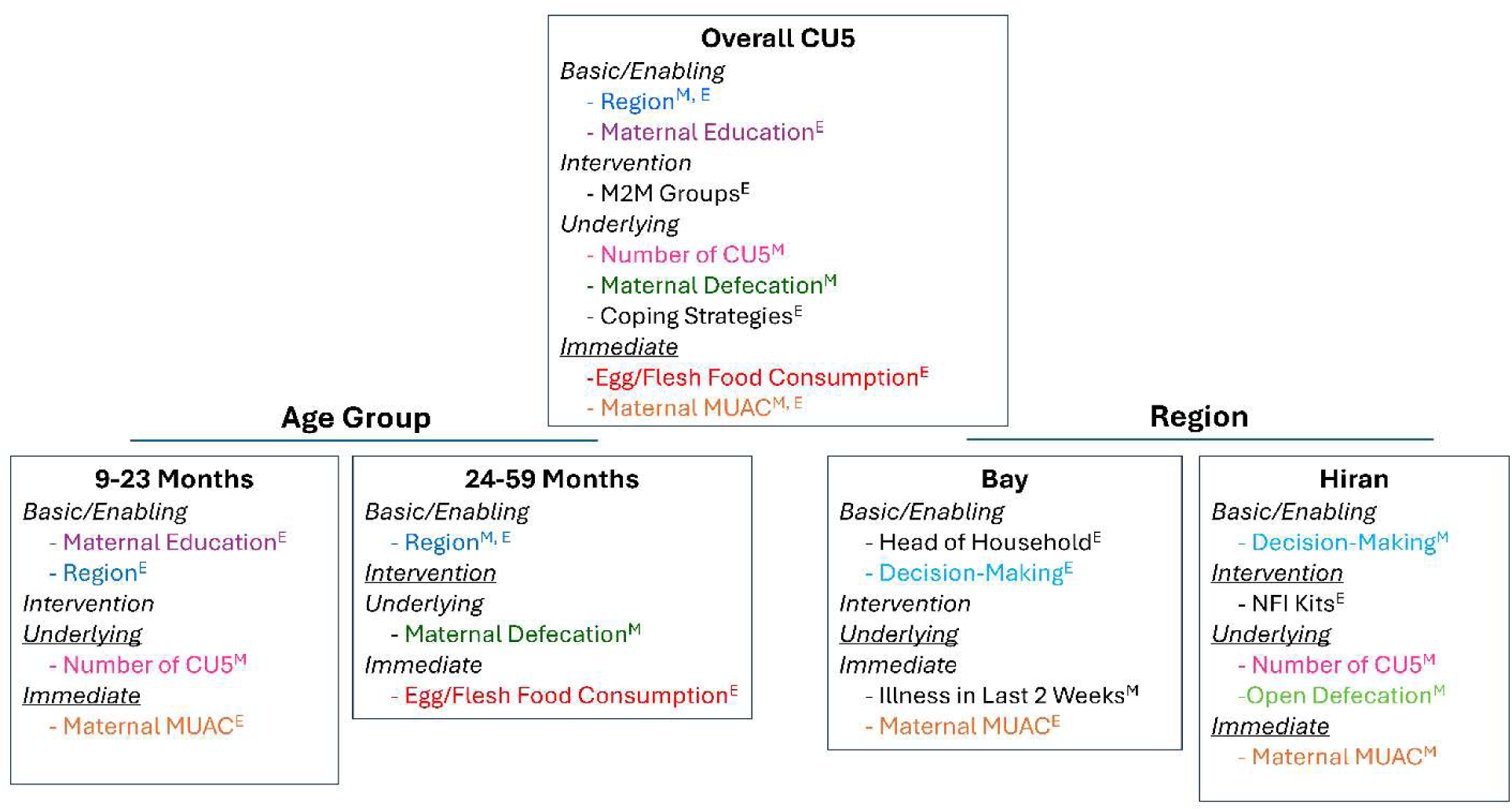
Drivers of CU5 WHZ in the Final Adjusted Models. *Color coding indicates that a variable was a statistically significant driver of child WHZ in multiple CU5 models. “M” superscript indicates the variable was a statistically significant driver in the given model at midline, “E” superscript indicates the variable was a statistically significant driver in the given model at endline, and “M,E” indicates the variable was a statistically significant driver in both the midline and endline final adjusted models. Note that this image depicts statistically significant drivers from the final adjusted models at the α = 0.05 level; additional drivers may have been marginally statistically significant or statistically significant in the unadjusted or basic confounder-adjusted models*.

At the basic/enabling level, household region was a statistically significant driver of WHZ at both midline and endline in the fully adjusted overall models. Children in Hiran had an average WHZ -0.66 SD (95% CI: -0.80, -0.52) and -0.58 SD (95% CI: -0.74, -0.43) lower than children in Bay at midline and endline, respectively (**Table 3**). This finding is consistent with results from the multivariable Poisson regression models where, at both midline and endline, children living in Hiran versus Bay had an increased relative risk of wasting. The education status of mothers was statistically significantly associated with WHZ at endline, with having no education corresponding to a -0.17 SD decrease in WHZ in the final overall adjusted model (**Table 3**, 95% CI: -0.33, -0.02).

**Table 3.**
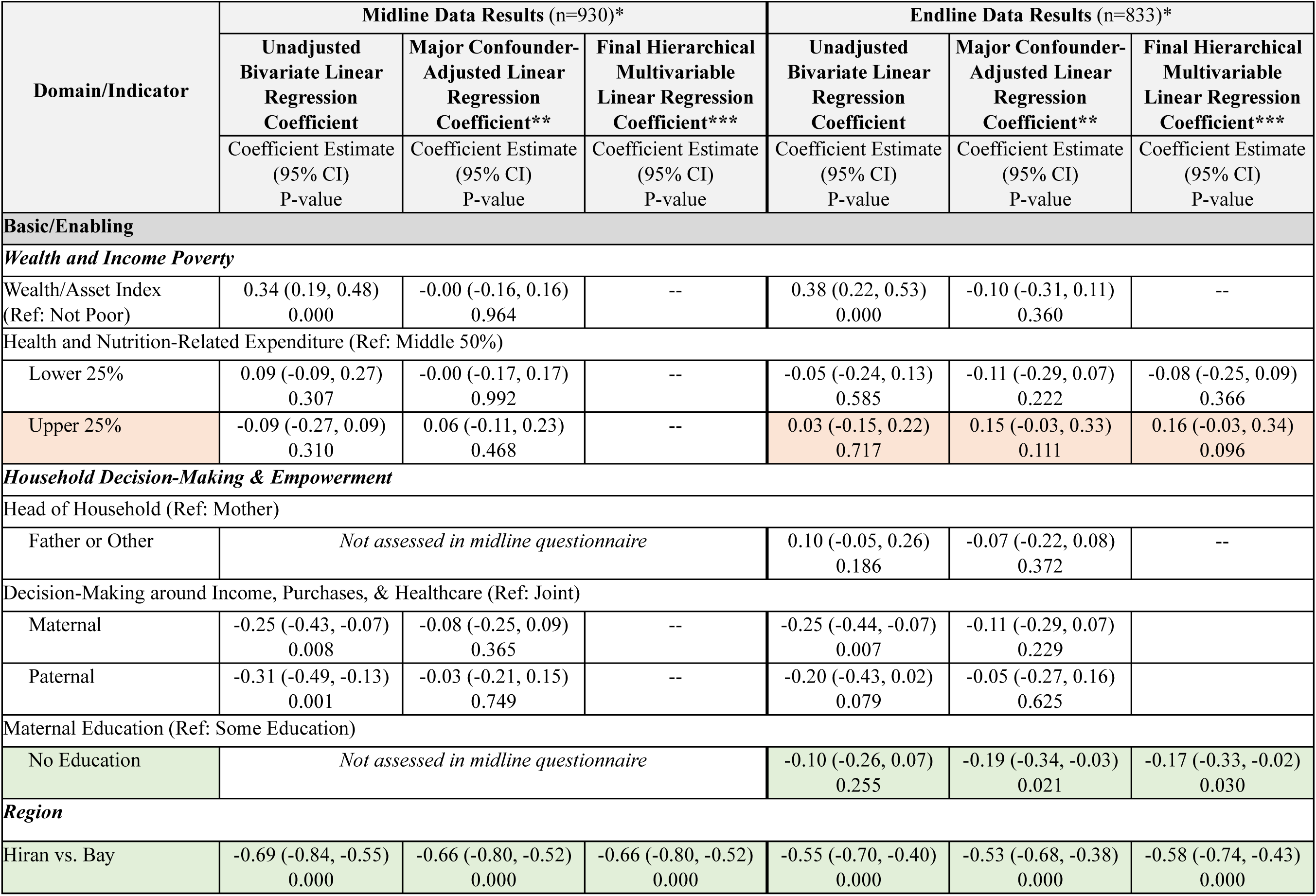
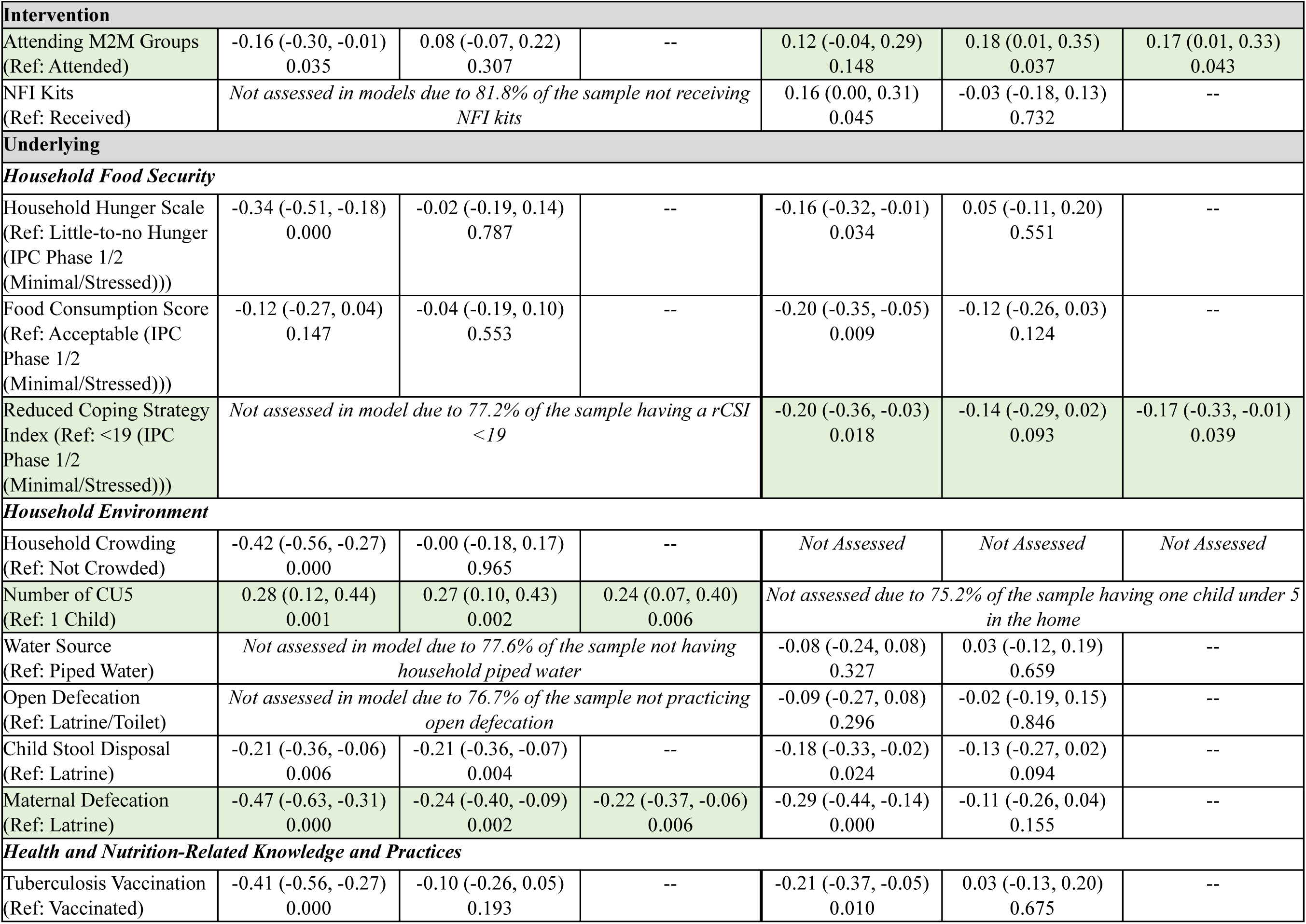
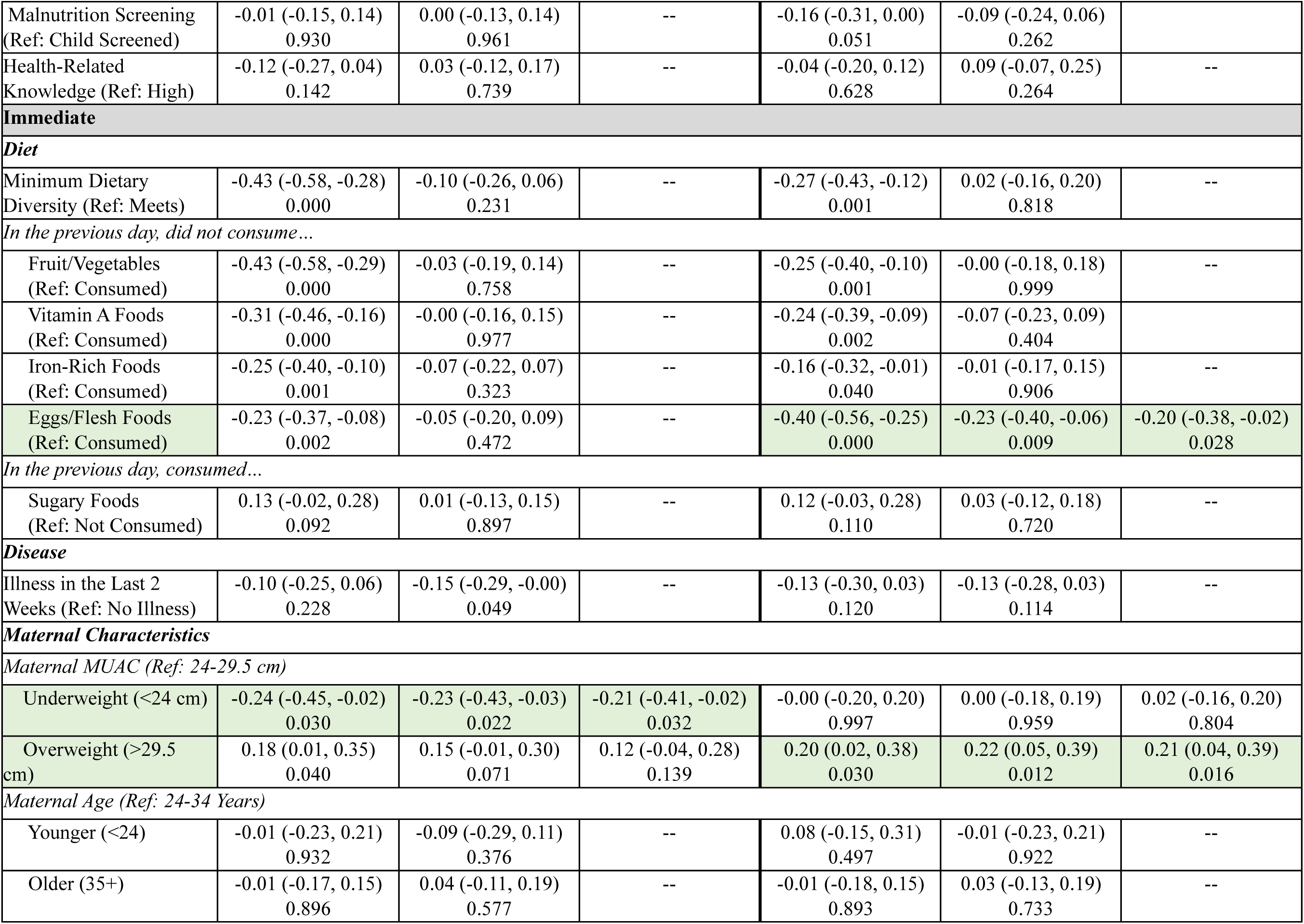
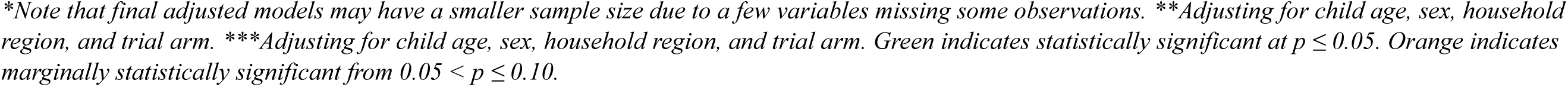
Drivers of CU5 Weight-for-Height Z-Score at Midline and Endline

| Domain/Indicator | Midline Data Results (n=930)* |  |  | Endline Data Results (n=833)* |  |  |
| --- | --- | --- | --- | --- | --- | --- |
|  | Unadjusted Bivariate Linear Regression Coefficient | Major Confounder-Adjusted Linear Regression Coefficient** | Final Hierarchical Multivariable Linear Regression Coefficient*** | Unadjusted Bivariate Linear Regression Coefficient | Major Confounder-Adjusted Linear Regression Coefficient** | Final Hierarchical Multivariable Linear Regression Coefficient*** |
|  | Coefficient Estimate (95% CI)<br>P-value | Coefficient Estimate (95% CI)<br>P-value | Coefficient Estimate (95% CI)<br>P-value | Coefficient Estimate (95% CI)<br>P-value | Coefficient Estimate (95% CI)<br>P-value | Coefficient Estimate (95% CI)<br>P-value |
| Basic/Enabling |  |  |  |  |  |  |
| Wealth and Income Poverty |  |  |  |  |  |  |
| Wealth/Asset Index (Ref: Not Poor) | 0.34 (0.19, 0.48)<br>0.000 | -0.00 (-0.16, 0.16)<br>0.964 | -- | 0.38 (0.22, 0.53)<br>0.000 | -0.10 (-0.31, 0.11)<br>0.360 | -- |
| Health and Nutrition-Related Expenditure (Ref: Middle 50%) |  |  |  |  |  |  |
| Lower 25% | 0.09 (-0.09, 0.27)<br>0.307 | -0.00 (-0.17, 0.17)<br>0.992 | -- | -0.05 (-0.24, 0.13)<br>0.585 | -0.11 (-0.29, 0.07)<br>0.222 | -0.08 (-0.25, 0.09)<br>0.366 |
| Upper 25% | -0.09 (-0.27, 0.09)<br>0.310 | 0.06 (-0.11, 0.23)<br>0.468 | -- | 0.03 (-0.15, 0.22)<br>0.717 | 0.15 (-0.03, 0.33)<br>0.111 | 0.16 (-0.03, 0.34)<br>0.096 |
| Household Decision-Making & Empowerment |  |  |  |  |  |  |
| Head of Household (Ref: Mother) |  |  |  |  |  |  |
| Father or Other | Not assessed in midline questionnaire |  |  | 0.10 (-0.05, 0.26)<br>0.186 | -0.07 (-0.22, 0.08)<br>0.372 | -- |
| Decision-Making around Income, Purchases, & Healthcare (Ref: Joint) |  |  |  |  |  |  |
| Maternal | -0.25 (-0.43, -0.07)<br>0.008 | -0.08 (-0.25, 0.09)<br>0.365 | -- | -0.25 (-0.44, -0.07)<br>0.007 | -0.11 (-0.29, 0.07)<br>0.229 |  |
| Paternal | -0.31 (-0.49, -0.13)<br>0.001 | -0.03 (-0.21, 0.15)<br>0.749 | -- | -0.20 (-0.43, 0.02)<br>0.079 | -0.05 (-0.27, 0.16)<br>0.625 |  |
| Maternal Education (Ref: Some Education) |  |  |  |  |  |  |
| No Education | Not assessed in midline questionnaire |  |  | -0.10 (-0.26, 0.07)<br>0.255 | -0.19 (-0.34, -0.03)<br>0.021 | -0.17 (-0.33, -0.02)<br>0.030 |
| Region |  |  |  |  |  |  |
| Hiran vs. Bay | -0.69 (-0.84, -0.55)<br>0.000 | -0.66 (-0.80, -0.52)<br>0.000 | -0.66 (-0.80, -0.52)<br>0.000 | -0.55 (-0.70, -0.40)<br>0.000 | -0.53 (-0.68, -0.38)<br>0.000 | -0.58 (-0.74, -0.43)<br>0.000 |

| Intervention |  |  |  |  |  |  |
| --- | --- | --- | --- | --- | --- | --- |
| Attending M2M Groups<br>(Ref: Attended) | -0.16 (-0.30, -0.01)<br>0.035 | 0.08 (-0.07, 0.22)<br>0.307 | -- | 0.12 (-0.04, 0.29)<br>0.148 | 0.18 (0.01, 0.35)<br>0.037 | 0.17 (0.01, 0.33)<br>0.043 |
| NFI Kits<br>(Ref: Received) | Not assessed in models due to 81.8% of the sample not receiving<br>NFI kits |  |  | 0.16 (0.00, 0.31)<br>0.045 | -0.03 (-0.18, 0.13)<br>0.732 | -- |
| Underlying |  |  |  |  |  |  |
| Household Food Security |  |  |  |  |  |  |
| Household Hunger Scale<br>(Ref: Little-to-no Hunger<br>(IPC Phase 1/2<br>(Minimal/Stressed)))) | -0.34 (-0.51, -0.18)<br>0.000 | -0.02 (-0.19, 0.14)<br>0.787 | -- | -0.16 (-0.32, -0.01)<br>0.034 | 0.05 (-0.11, 0.20)<br>0.551 | -- |
| Food Consumption Score<br>(Ref: Acceptable (IPC<br>Phase 1/2<br>(Minimal/Stressed)))) | -0.12 (-0.27, 0.04)<br>0.147 | -0.04 (-0.19, 0.10)<br>0.553 | -- | -0.20 (-0.35, -0.05)<br>0.009 | -0.12 (-0.26, 0.03)<br>0.124 | -- |
| Reduced Coping Strategy<br>Index (Ref: <19 (IPC<br>Phase 1/2<br>(Minimal/Stressed)))) | Not assessed in model due to 77.2% of the sample having a rCSI<br><19 |  |  | -0.20 (-0.36, -0.03)<br>0.018 | -0.14 (-0.29, 0.02)<br>0.093 | -0.17 (-0.33, -0.01)<br>0.039 |
| Household Environment |  |  |  |  |  |  |
| Household Crowding<br>(Ref: Not Crowded) | -0.42 (-0.56, -0.27)<br>0.000 | -0.00 (-0.18, 0.17)<br>0.965 | -- | Not Assessed | Not Assessed | Not Assessed |
| Number of CU5<br>(Ref: 1 Child) | 0.28 (0.12, 0.44)<br>0.001 | 0.27 (0.10, 0.43)<br>0.002 | 0.24 (0.07, 0.40)<br>0.006 | Not assessed due to 75.2% of the sample having one child under 5<br>in the home |  |  |
| Water Source<br>(Ref: Piped Water) | Not assessed in model due to 77.6% of the sample not having<br>household piped water |  |  | -0.08 (-0.24, 0.08)<br>0.327 | 0.03 (-0.12, 0.19)<br>0.659 | -- |
| Open Defecation<br>(Ref: Latrine/Toilet) | Not assessed in model due to 76.7% of the sample not practicing<br>open defecation |  |  | -0.09 (-0.27, 0.08)<br>0.296 | -0.02 (-0.19, 0.15)<br>0.846 | -- |
| Child Stool Disposal<br>(Ref: Latrine) | -0.21 (-0.36, -0.06)<br>0.006 | -0.21 (-0.36, -0.07)<br>0.004 | -- | -0.18 (-0.33, -0.02)<br>0.024 | -0.13 (-0.27, 0.02)<br>0.094 | -- |
| Maternal Defecation<br>(Ref: Latrine) | -0.47 (-0.63, -0.31)<br>0.000 | -0.24 (-0.40, -0.09)<br>0.002 | -0.22 (-0.37, -0.06)<br>0.006 | -0.29 (-0.44, -0.14)<br>0.000 | -0.11 (-0.26, 0.04)<br>0.155 | -- |
| Health and Nutrition-Related Knowledge and Practices |  |  |  |  |  |  |
| Tuberculosis Vaccination<br>(Ref: Vaccinated) | -0.41 (-0.56, -0.27)<br>0.000 | -0.10 (-0.26, 0.05)<br>0.193 | -- | -0.21 (-0.37, -0.05)<br>0.010 | 0.03 (-0.13, 0.20)<br>0.675 | -- |
| Malnutrition Screening<br>(Ref: Child Screened) | -0.01 (-0.15, 0.14)<br>0.930 | 0.00 (-0.13, 0.14)<br>0.961 | -- | -0.16 (-0.31, 0.00)<br>0.051 | -0.09 (-0.24, 0.06)<br>0.262 |  |
| Health-Related<br>Knowledge (Ref: High) | -0.12 (-0.27, 0.04)<br>0.142 | 0.03 (-0.12, 0.17)<br>0.739 | -- | -0.04 (-0.20, 0.12)<br>0.628 | 0.09 (-0.07, 0.25)<br>0.264 | -- |
| **Immediate** |  |  |  |  |  |  |
| **Diet** |  |  |  |  |  |  |
| Minimum Dietary<br>Diversity (Ref: Meets) | -0.43 (-0.58, -0.28)<br>0.000 | -0.10 (-0.26, 0.06)<br>0.231 | -- | -0.27 (-0.43, -0.12)<br>0.001 | 0.02 (-0.16, 0.20)<br>0.818 | -- |
| *In the previous day, did not consume...* |  |  |  |  |  |  |
| Fruit/Vegetables<br>(Ref: Consumed) | -0.43 (-0.58, -0.29)<br>0.000 | -0.03 (-0.19, 0.14)<br>0.758 | -- | -0.25 (-0.40, -0.10)<br>0.001 | -0.00 (-0.18, 0.18)<br>0.999 | -- |
| Vitamin A Foods<br>(Ref: Consumed) | -0.31 (-0.46, -0.16)<br>0.000 | -0.00 (-0.16, 0.15)<br>0.977 | -- | -0.24 (-0.39, -0.09)<br>0.002 | -0.07 (-0.23, 0.09)<br>0.404 |  |
| Iron-Rich Foods<br>(Ref: Consumed) | -0.25 (-0.40, -0.10)<br>0.001 | -0.07 (-0.22, 0.07)<br>0.323 | -- | -0.16 (-0.32, -0.01)<br>0.040 | -0.01 (-0.17, 0.15)<br>0.906 | -- |
| Eggs/Flesh Foods<br>(Ref: Consumed) | -0.23 (-0.37, -0.08)<br>0.002 | -0.05 (-0.20, 0.09)<br>0.472 | -- | -0.40 (-0.56, -0.25)<br>0.000 | -0.23 (-0.40, -0.06)<br>0.009 | -0.20 (-0.38, -0.02)<br>0.028 |
| *In the previous day, consumed...* |  |  |  |  |  |  |
| Sugary Foods<br>(Ref: Not Consumed) | 0.13 (-0.02, 0.28)<br>0.092 | 0.01 (-0.13, 0.15)<br>0.897 | -- | 0.12 (-0.03, 0.28)<br>0.110 | 0.03 (-0.12, 0.18)<br>0.720 | -- |
| **Disease** |  |  |  |  |  |  |
| Illness in the Last 2<br>Weeks (Ref: No Illness) | -0.10 (-0.25, 0.06)<br>0.228 | -0.15 (-0.29, -0.00)<br>0.049 | -- | -0.13 (-0.30, 0.03)<br>0.120 | -0.13 (-0.28, 0.03)<br>0.114 | -- |
| **Maternal Characteristics** |  |  |  |  |  |  |
| *Maternal MUAC (Ref: 24-29.5 cm)* |  |  |  |  |  |  |
| Underweight (<24 cm) | -0.24 (-0.45, -0.02)<br>0.030 | -0.23 (-0.43, -0.03)<br>0.022 | -0.21 (-0.41, -0.02)<br>0.032 | -0.00 (-0.20, 0.20)<br>0.997 | 0.00 (-0.18, 0.19)<br>0.959 | 0.02 (-0.16, 0.20)<br>0.804 |
| Overweight (>29.5<br>cm) | 0.18 (0.01, 0.35)<br>0.040 | 0.15 (-0.01, 0.30)<br>0.071 | 0.12 (-0.04, 0.28)<br>0.139 | 0.20 (0.02, 0.38)<br>0.030 | 0.22 (0.05, 0.39)<br>0.012 | 0.21 (0.04, 0.39)<br>0.016 |
| *Maternal Age (Ref: 24-34 Years)* |  |  |  |  |  |  |
| Younger (<24) | -0.01 (-0.23, 0.21)<br>0.932 | -0.09 (-0.29, 0.11)<br>0.376 | -- | 0.08 (-0.15, 0.31)<br>0.497 | -0.01 (-0.23, 0.21)<br>0.922 | -- |
| Older (35+) | -0.01 (-0.17, 0.15)<br>0.896 | 0.04 (-0.11, 0.19)<br>0.577 | -- | -0.01 (-0.18, 0.15)<br>0.893 | 0.03 (-0.13, 0.19)<br>0.733 | -- |

*\*Note that final adjusted models may have a smaller sample size due to a few variables missing some observations. \*\*Adjusting for child age, sex, household region, and trial arm. \*\*\*Adjusting for child age, sex, household region, and trial arm. Green indicates statistically significant at $p \leq 0.05$ . Orange indicates marginally statistically significant from $0.05 < p \leq 0.10$ .*

At the intervention level, not attending mother-to-mother (M2M) support groups was positively associated with WHZ in the endline overall model (**Table 3**, Coefficient= 0.17, 95% CI: 0.01, 0.33), likely due to reverse causality.

At the underlying level, having more than one CU5 in the household was protective of WHZ in the overall model at midline (**Table 3**, Coefficient= 0.24, 95% CI: 0.07, 0.40). Maternal defecation in an open area versus a latrine adversely affected WHZ in the overall midline final adjusted model (**Table 3**, Coefficient= -0.22, 95% CI: -0.37, -0.06). Similarly, household disposal of children’s stool in an open area versus a latrine was associated with a higher risk of wasting in the Poisson regression endline model. Regarding food insecurity, having a rCSI score ≥19 (crisis/emergency/famine IPC phase) was negatively associated with WHZ (**Table 3**, Coefficient= -0.17, 95% CI: -0.33, -0.01) in the endline overall final adjusted model.

At the immediate level, the mother being underweight adversely affected WHZ in the midline overall model (**Table 3**, Coefficient= -0.21, 95% CI: -0.41, -0.02), and, similarly, the mother being overweight was protective of WHZ in the overall endline model (**Table 3**, Coefficient= 0.21, 95% CI: 0.04, 0.39). The child not consuming eggs and/or flesh foods the previous day was associated with a lower WHZ in the endline overall adjusted model (**Table 3**, Coefficient= -0.20, 95% CI: -0.38, -0.02), demonstrating a protective effect of protein-rich food consumption.

When stratifying by child age, no new drivers of WHZ were identified. Rather, household region remained a driver in the endline 9–23-month model and in both the midline and endline 24– 59-month models, maternal education was identified as a driver in the endline 9-23 month model, and having more than one CU5 was again protective of WHZ in the midline 9-23 month model. Maternal defecation practices and egg and/or flesh foods consumption were associated with WHZ in the midline and endline 24–59-month models, respectively, and the mother being overweight was again protective of WHZ in the endline 9–23-month model.

When stratifying by region, some drivers consistent with the overall models were identified while a few new statistically significant drivers emerged. At midline in Hiran, having more than one CU5 in the household remained as a protective driver of WHZ and having an underweight mother was again correlated with lower WHZ. Similarly, in Bay at endline, the mother being overweight again appeared as being protective of WHZ. On the other hand, at midline in Hiran and at endline in Bay, maternal decision-making, as opposed to joint decision-making, was correlated with a lower WHZ, providing evidence of joint decision-making being protective of child WHZ. In Hiran, household open defecation was negatively correlated with WHZ at midline, while not receiving a non-food item (NFI) kit (kits were distributed to the most vulnerable households) was protective at endline. In Bay, recent child illness adversely affected WHZ at midline and living in a father- or other-headed household was associated with a lower WHZ in comparison to maternal-headed households at endline.

### Drivers of Mothers’ Wasting

While additional drivers may have been statistically significant in the unadjusted or basic confounder-adjusted models, statistically significant findings in the final hierarchical adjusted models are presented here (see **Tables S14** and **S15**).

Maternal household decision-making was associated with wasting at midline, with mothers living in maternal-decision-making households having a lower relative risk of wasting compared to joint decision-making households. Not attending M2M groups was statistically significantly associated with wasting at endline. At the underlying level, having an unacceptable FCS was associated with an increased relative risk of wasting in comparison to households with an acceptable FCS at midline. Relatedly, mothers in households with moderate-severe hunger according to the HHS had an increased relative risk of wasting in comparison to households with little-to-no hunger at midline. Disposing of children’s stool in an open area was associated with a higher risk of wasting at midline, with household practice of open defecation associated with a higher risk of wasting at endline. At the immediate level, age was statistically significantly associated with wasting at midline, with older mothers (35+) having a lower relative risk of wasting in comparison to mothers aged 24-34 years.

## Discussion

To the authors’ knowledge, this analysis is the first study to explore the drivers of wasting among both CU5 and their mothers in two high wasting burden regions of Somalia. The burden of child wasting differed greatly by region, with Hiran having approximately a 3-4 times higher burden of child wasting compared to Bay, and region appeared as a driver of child WHZ in five out of six relevant child models (excluding the region-stratified models). The variation in wasting across regions justified the need to stratify the analyses by region to better understand the region-specific factors contributing to wasting. While maternal MUAC and household decision-making were statistically significant drivers in both regions, additional unique factors were identified in each region. When the analyses were stratified by child age, the only common driver between children aged 9-23 months and those aged 24-59 months was region. However, additional factors influencing WHZ were identified for each age group, showing that the most significant drivers of child nutrition outcomes differed depending on the child’s age. For example, consumption of egg and/or flesh foods was retained as a driver of WHZ among children aged 24-59 months at endline, but not among younger children at endline, which may be due to animal-based protein foods perhaps being a more important food group for children as they get older, with infants and children under 2 being more reliant on breastmilk. Regarding seasonality, multiple drivers of WHZ differed between midline (Xagaa dry season/Deyr rainy season transition) and endline (Jilaal dry season), with only household region, household decision-making, and maternal MUAC appearing as statistically significant drivers at both timepoints across different models. Previous work has shown that seasonality is a significant predictor of malnutrition among CU5 in Somalia, supporting that differences in drivers may be observed by season.[10,12,13] Further supporting this finding, none of the drivers of mothers’ wasting were the same between midline and endline. Taken together, these findings highlight the importance of recognizing the context-specific nature of drivers of child and maternal wasting in program efforts, understanding that the most significant drivers in a given context may vary by sub-population, region, and season. However, these differences in identified drivers by region should be interpreted with caution. Across different models, indicators were excluded in certain analyses if there was high imbalance of the indicator in the analysis sample (see data tables). As a result, the regional, seasonal, and child age differences in drivers may not fully account for all relevant factors.

Multiple characteristics of maternal empowerment were associated with child nutrition status. The mother having no formal education and the father being the head of the household were associated with lower WHZ in different stratified CU5 models. These findings are consistent with previous work in Ethiopia, where maternal illiteracy and lack of education were risk factors for wasting among children aged 6-59 months.[42–44] Regarding household decision-making, different results were found between children and mothers. Not having joint household decision-making between both parents was associated with lower child WHZ, and these findings are consistent with work done in Ethiopia where children in households with maternal-only or paternal-only decision-making were at an increased risk of wasting compared to joint household decision-making.[44] On the other hand, maternal decision-making was found to be a protective factor against maternal wasting compared to joint household decision-making, aligning with findings from studies in Ethiopia where household decision-making and control were predictive of maternal nutrition status.[44–47] As one hypothesis for these results, husbands’ involvement in joint decision-making may reflect more involvement in child care and therefore be beneficial for a child’s nutrition; for example, a study in Nepal found that joint decision-making reflected great husband involvement in pregnancy care.[48,49] A possible explanation for the protective effect of maternal-only decision-making on maternal wasting is that mothers can prioritize their own health more effectively, aligning with studies that link greater autonomy to better health service utilization.[46,50]

Not receiving a NFI kit was protective of WHZ. As the most vulnerable households were selected to receive NFI kits, this result supports that those who did not receive an NFI kit were less vulnerable and more protected against wasting. Additionally, the mother not attending M2M groups showed a protective relationship with both child WHZ and maternal wasting at endline. However, this finding does not suggest that M2M groups were harmful, but rather, as this variable is an intervention component measured in a cross-sectional survey, the phenomenon of reverse causality is likely, where those who receive intervention components (in this case, attending M2M groups) may be the most vulnerable and already have worse nutrition outcomes.[51–56] Therefore, it is not that attending the groups causes poor nutrition outcomes, but rather, it is likely that individuals with poor outcomes are more likely to attend the groups.

Multiple sanitation indicators were found to be drivers of wasting, consistent with previously identified causes of wasting.[31,57,58] Poor sanitation practices, such as open defecation and improper disposal of stool, were identified risk factors for both maternal and child wasting. These practices likely increase the risk of infection and illness, which, as this study and others in Somalia have shown, is a key driver of wasting.[10–13,59] These results emphasize the importance of improving sanitation practices, such as by improving infrastructure to increase the accessibility of latrines among households.

Food security is an established risk factor for wasting, and household food security has previously been identified as a significant determinant of nutritional status among pregnant women in Ethiopia in multiple studies.[31,45,57,60–62] Consistent with previous literature, both an unacceptable food consumption score and moderate-to-severe household hunger were risk factors for mothers’ wasting in this analysis.[31,45,57,60–63] Similarly, at endline, a higher rCSI score, corresponding to crisis/extreme/famine levels of food security, was correlated with lower child WHZ. These findings emphasize promoting household food security to support the nutrition status of children and mothers in this setting, such as by ensuring access to local markets equipped with affordable and nutritious foods.

Having more than one CU5 in a household was protective of WHZ in multiple models, contrary to analyses from Somalia national survey data, where having more CU5 in a household was correlated with a higher risk of child wasting.[10,12,13] However, data from Uganda has shown that having siblings is protective of child stunting, demonstrating that having multiple children may be protective of child nutrition status.[64]

Maternal MUAC was the most frequently appearing statistically significant driver of WHZ throughout different models, and maternal malnutrition is a cited risk factor for child wasting, emphasizing the close relationship between maternal characteristics and child nutrition outcomes.[57] This study’s results are consistent with previous work in Somalia, where a higher mother’s MUAC was correlated with decreased wasting among children aged 6-59 months.[11–13] The association between maternal MUAC and child WHZ demonstrates the intergenerational and connected aspects of maternal and child nutrition status and supports designing policies and programs to protect the nutrition status of mothers.[57,65,66]

Consumption of egg and/or flesh foods was protective of WHZ. Animal-based foods reflect improved nutrient intake for ideal child growth[67], and these findings are consistent with previous work in Somalia and Ethiopia where protein and animal source foods were important for the nutrition status of CU5.[10–13] These findings highlight the importance of ensuring that high protein, animal-based foods are available in local markets and that households have the financial means to afford such foods.

Limitations of this analysis include the potential for recall bias, and the potential effect of unmeasured confounders. Additional limitations include data for some variables only being available at one time point, not being able to include some indicators in specific models due to high imbalance in the analysis sample, and the questionnaire not including questions about maternal diet and disease, limiting the ability to analyze important immediate determinants of maternal nutrition status. Nevertheless, strengths of this study include the ability to analyze a wide breadth of indicators spanning the entire conceptual framework of maternal and child nutrition, to study the relationship between maternal and child nutrition, and to utilize a hierarchical, multivariable model-building approach to arrive at fully adjusted models that minimize the effects of confounding. Additionally, while the present study was a cross-sectional analysis, the data was from a prospective cohort study and therefore allowed for comparing the drivers of wasting at two different time points corresponding to different seasons, and stratified models were used to study associations within strata of key confounding variables.[68] While causality cannot be established from a cross-sectional analysis, covariates were selected according to biological plausibility and coherence with existing knowledge, and the statistically significant findings from the fully adjusted models demonstrate the strength of association of identified drivers.[68–70] Although cross-sectional analysis does not allow for examining direct temporal relationships, wasting is a dynamic condition that can change quickly. By analyzing factors that were present before assessing participants’ nutrition status—such as illness in the past two weeks or vaccination status—some insights into exposure history can still be inferred.[70]

Findings are generalizable to similar such humanitarian contexts (such as conflict- and climate-affected contexts) in Somalia and other countries. Future analyses should explore the drivers of maternal and child wasting in other high wasting burden areas of Somalia as well as during different seasons to construct a more complete picture of how the most significant drivers of wasting may fluctuate throughout a calendar year. Future research should also examine characteristics of maternal diet and disease as potential drivers of mothers’ wasting and further study gender dynamics in this context and the implications for household resources, food and nutrition practices, and health-seeking behaviors.

## Conclusion

To address the high burden of wasting in Somalia, it is important to explore the most significant drivers of maternal and child wasting. This study highlights regional, age, and seasonal variation in the drivers of wasting, underscoring the importance of context-specific approaches to addressing malnutrition. Key drivers, such as maternal MUAC and household decision-making, were consistent across regions, while others, including food security and maternal empowerment, varied by region, season, and age group.

The findings emphasize the need for tailored interventions that consider local factors like household food security and sanitation practices, seasonality, and gender dynamics. The study also highlights the interconnectedness of maternal and child nutritional status, reinforcing the importance of maternal health in improving child nutrition outcomes.

Future research should further explore the nature and implications of gender dynamics in this setting on maternal and child nutrition and should characterize the drivers of wasting during other seasons and in other high-wasting burden areas of Somalia.

These findings contribute to a growing body of literature on the complex, multifactorial drivers of wasting and underscore the importance of designing multisectoral interventions that address the immediate, underlying, and basic causes of malnutrition.

## Ethics

All participants were informed of the study purpose, the use of the collected data, and any potential risks to participation. Individuals that agreed to participate signed an informed consent form. Approval for this study was obtained from the Johns Hopkins Bloomberg School of Public Health Institutional Review Board (protocol code24476), the Somalia Ministry of Health and Human Services (XAG/203/23), and the Ethics Review Committee at Save the Children International.

## Data Availability

This data was collected on a vulnerable population linked to a large-scale cash assistance program in Somalia. The data may be available upon request with permission from local study investigators.

## Authorship Contributions

Conceptualization (S.G., N.A., S.W., S.G., S.A.M.); data collection (S.A.M., S.A., M.A.N., A.M.M., M.O., M.B.M., A.A., A.M.); data cleaning and analysis (S.G., N.A., K.K.A., S.G.); draft manuscript preparation (S.G., N.A., S.W., S.G.); critical review and editing of manuscript (S.G., S.W., K.K.A., S.G., S.A.M., S.A., Q.K., M.A.N., A.M.M., M.B.M., M.T., N.A.). All authors reviewed and approved the final version.

## Acknowledgements

The authors acknowledge the important contributions of all study participants, the Somalia Ministries of Health, and the data collection team.

