## Supplemental Files for "Assessing the Drivers of Wasting among Children Under 5 and Their Mothers in The Bay and Hiran Regions of Somalia"

#### Supplementary Materials

#### Table of Contents

**Appendix S1.** Technical Appendix

**Text S1:** Technical Definition of Wasting
**Text S2:** Somalia CashPlus for Nutrition cRCT Overview
**Text S3:** Study Setting and Context
**Table S1.** Independent Variables Tested as Drivers of CU5 and Mothers' Wasting
**Table S2:** Definitions of Key Indicators
**Text S4:** Construction of Different Indicators
**Figure S1.** Mapping of Variables Tested in the Mothers' Models
**Text S5:** Defining the Study Sample
**Text S6:** Modeling Strategy

**Appendix S2.** Results Appendix

**Table S3.** Additional Child, Mother, and Household Characteristics
**Table S4.** Midline Overall CU5 Model: Drivers of Wasting
**Table S5.** Endline Overall CU5 Model: Drivers of Wasting
**Table S6.** Midline 9-23 Months Model: Drivers of Weight-for-Height Z-Score
**Table S7.** Midline 24-59 Months Model: Drivers of Weight-for-Height Z-Score
**Table S8.** Midline Hiran Region CU5 Model: Drivers of Weight-for-Height Z-Score
**Table S9.** Midline Bay Region CU5 Model: Drivers of Weight-for-Height Z-Score
**Table S10.** Endline 9-23 Month CU5 Model: Drivers of Weight-for-Height Z-Score
**Table S11.** Endline 24-59 Month CU5 Model: Drivers of Weight-for-Height Z-Score
**Table S12.** Endline Hiran Region CU5 Model: Drivers of Weight-for-Height Z-Score
**Table S13.** Endline Bay CU5 Model: Drivers of Weight-for-Height Z-Score
**Figure S2.** Drivers of Mothers' Wasting
**Table S14.** Midline Overall Maternal MUAC Model: Drivers of Wasting
**Table S15.** Endline Overall Maternal MUAC Model: Drivers of Wasting

### **Appendix S1.** Technical Appendix

**Text S1:** Technical Definition of Wasting

Wasting among children is defined by a weight-for-height Z-score (WHZ) measurement < -2 standard deviations (SD) from the 2006 WHO Child Growth Standards median and is further characterized as either moderate (< -2 but ≥ -3 SD) or severe (< -3 SD).[1–4] Wasting among mothers is defined as a mid-upper arm circumference (MUAC) of less than 23 centimeters.[5–7]

**Text S2:** Somalia CashPlus for Nutrition cRCT Overview

Data for this study comes from a prospective, mixed methods cluster randomized controlled trial (cRCT) to evaluate the effectiveness of a CashPlus for Nutrition program implemented by Save the Children (SC) in the Bay and Hiran regions of Somalia from June-November 2023.[8,9] The specific study sites were the capitals of these regions, Baidoa (Bay) and Beledweyne (Hiran). The CashPlus for Nutrition project was aimed at preventing acute malnutrition among CU5 and their mothers through various combinations of cash and social and behavior change communication (SBCC) interventions.[8,9]

The cRCT lasted for six months, and villages were randomized to three study arms: 1) cash only, 2) cash plus SBCC, and 3) cash plus additional cash top-up.[8,9] Participants were mothers of children under 5 (CU5) enrolled in the CashPlus for Nutrition program who were contacted and recruited for participation in the trial. Participants received cash monthly via a mobile transfer, with participants in Bay receiving an amount equivalent to 90 US dollars (USD) and participants in Hiran receiving an amount equivalent to 70 USD, according to the minimum expenditure basket.[8,9] Those randomized to the cash plus additional cash top-up trial arm received an additional 35 USD per month.[8,9] Participants randomized to the cash plus SBCC trial arm received community-level promotion campaigns, individual sessions, and bi-monthly mother-to-mother (M2M) group sessions around health and nutrition-related issues.[8,9] Additional details of the cRCT are published elsewhere.[8,9] For the purposes of this study, the cRCT trial arm was adjusted for in the analysis to control for any effect of the interventions.

**Text S3:** Study Setting & Context

Somalia is found in the east of the Horn of Africa and is characterized by a tropical, mediterranean climate.[10,11] Somalia's calendar year is characterized by four seasons: two rainy seasons and two dry seasons.[10] Jilaal dry season occurs from December to March, followed by the Gu rainy season that occurs in the latter half of March through June.[12] The Gu rainy season accounts for the majority of the country's rainfall, at about 75% of total rainfall.[12] The Xagaa dry season then takes place from July through September, with some rainfall occurring in certain parts of the country.[10,12] Finally, the Deyr rainy season occurs from September through November, accounting for about a quarter of the country's annual rainfall.[10,12] Over the years, climate change has disrupted typical weather patterns, resulting in repeated droughts and flooding throughout Somalia.[10]

Climate-related issues (droughts, flooding) and conflict have contributed to issues of food insecurity and displacement, causing Somalia to be characterized as a "complex emergency."[13]

It is estimated that nearly six million individuals in Somalia will require humanitarian assistance in 2025, and Somalia is estimated to have 3.3 million internally displaced people (IDPs).[13] Approximately 4.4 million individuals were also expected to experience acute food insecurity from October-December 2024.[13]

This study took place in the Bay and Hiran regions of Somalia, found in the southwest region of the country.[14–16] Recent reports estimate a population of approximately 1.06 million people in Bay and roughly 0.4 million people in Hiran, with the majority of individuals in both regions being rural/nomadic, but with Hiran having a higher percentage of individuals living in urban areas compared to Bay.[17–21] Consistent with the situation seen throughout the country, flooding, droughts, and conflict have threatened the livelihoods of individuals in Bay and Hiran.[15–17,22–24] As of December 2023, more than 100,000 people in both Bay and Hiran regions (separately) were reported to have been affected by the flooding that began in Somalia in October 2023.[22]

Frequent droughts and flooding have contributed to both displacement from these regions as well as arrivals of IDPs to these regions throughout the years.[15–17,23,24] For example, while over a quarter of a million individuals were displaced from Hiran from July 2021-November 2022, close to half a million newly displaced people arrived in Hiran in 2023.[15,16,24,25] Similar trends are seen in Bay, where thousands of individuals have been both displaced from and arrived in the Bay region as a result of such instability.[15,17,23–25] Baidoa city in Bay is home to approximately 25% of Somalia's IDP sites, with 0.75 million Bay residents experiencing crisis or emergency levels of food insecurity from January-March 2023.[17,24]

**Table S1.** Independent Variables Tested as Drivers of CU5 and Mothers' Wasting

| **Area of Interest** | **Independent Variable** | **Categories (Coding)** | **Reference Category** |
|---|---|---|---|
| Location | Region | Bay (0), Hiran (1) | Bay |
| Wealth and Income Poverty | Wealth/Asset Index | Not Poor (0), Poor (1) | Not Poor |
| | Health- and Nutrition-Related Monthly Expenditure | Middle 50% (0), Lower 25% (1), Upper 25% (2) | Middle 50% |
| Displacement | Displacement in the Last 3 Months | Not Displaced (0), Displaced (1) | Not Displaced |
| Household Decision-Making and Women's Empowerment | Head of Household | Mother (0), Father/Other (1) | Mother |
| | Household Decision-Making | Joint (0), Maternal (1), Paternal (2) | Joint |
| | Maternal Education* | No Education (0), Some Education (1) | Some Education |
| Intervention Components | Mother-to-Mother Groups | Attended (0), Did not Attend (1) | Attended |
| | Non-Food Item Kits** | Received (0), Did not Receive (1) | Received |

| | | | |
|---|---|---|---|
| Household Food Security | Household Hunger Scale | Little-to-No Hunger (0), Moderate-to-Severe Hunger (1) | Little-to-No Hunger (IPC Phase 1/2 (Minimal/ Stressed)) |
| | Food Consumption Score | Acceptable (36+) (0), Unacceptable (<36) (1) | Acceptable (IPC Phase 1/2 (Minimal/ Stressed) |
| | Reduced Coping Strategy Index | Score of <19 (0), Score of ≥19 (1) | Score of <19 (IPC Phase 1/2 (Minimal/ Stressed) |
| Household Environment | Number of CU5 | 1 Child (0), 2+ Children (1) | 1 Child |
| | Household Crowding | Not Crowded (0), Crowded (1) | Not Crowded |
| | Water Treatment | Treats (0), Does not Treat (1) | Treats |
| | Water Source | Piped (0), Not Piped (1) | Piped |
| | Household Defecation | Latrine/Toilet (0), Open Defecation (1) | Latrine/Toilet |
| | Child Stool Disposal | Latrine/Toilet (0), Open Area (1) | Latrine/Toilet |
| | Maternal Defecation Practices | Latrine/Toilet (0), Open Area (1) | Latrine/Toilet |
| Health- and Nutrition-Related Knowledge and Practices | Tuberculosis Vaccination*** | Vaccinated (0), Not Vaccinated (1) | Vaccinated |
| | Child Malnutrition Screening*** | Child Screened (0), Child Not Screened (1) | Child Screened |
| | Maternal Malnutrition Screening**** | Mother Screened (0), Mother Not Screened (1) | Mother Screened |
| | Health-Related Knowledge | High (0), Low-to-Moderate (1) | High |
| Child Diet*** | Ever Breastfed | Breastfed (0), Not Breastfed (1) | Breastfed |
| | Minimum Dietary Diversity | Meets (0), Does not Meet (1) | Meets |
| | Fruit/Vegetable Consumption | Consumed (0), Did not Consume (1) | Consumed |
| | Vitamin A-Rich Food Consumption | Consumed (0), Did not Consume (1) | Consumed |

| | | | |
|---|---|---|---|
| | Iron-Rich Food Consumption | Consumed (0), Did not Consume (1) | Consumed |
| | Egg and/or Flesh-Food Consumption | Consumed (0), Did not Consume (1) | Consumed |
| | Sugary Food Consumption | Did not Consume (0), Consumed (1) | Did not Consume |
| Child Disease*** | Illness in the Last 2 Weeks | Not Ill (0), Ill (1) | Not Ill |
| Maternal Characteristics | Maternal MUAC*** | Normal (24-29.5 cm) (0), Underweight (<24 cm) (1), Overweight (≥ 29.5 cm) (2) | Normal |
| | Maternal Age | Middle (24-34) (0), Younger (<24) (1), Older (35+) (2) | Middle |

*For maternal education, "some education" included madrasa, primary school, secondary school, and higher education. **The most vulnerable households were selected to receive WASH non-food item/hygiene kits according to a set of criteria determined by Save the Children. ***These variables were only tested in the CU5 models. ****This variable was only tested in the mothers' models.

**Table S2:** Definitions of Key Indicators

| ***Child-Related Key Indicators*** | | |
|---|---|---|
| **Indicator** | **Numerator** | **Denominator** |
| Child Illness in the Past 2 Weeks | Children in the study sample having diarrhea, fever, and/or a cough in the 2 weeks preceding the survey | Children in the study sample* |
| Tuberculosis Vaccination | Children in the study sample ever having received the tuberculosis vaccination (BCG against TB) | Children in the study sample |
| Child Ever Breastfed | Children in the study sample where the mother reported that the child has ever been breastfed[26] | Children in the study sample |
| Minimum Dietary Diversity 7 (MDDC7) | Children in the study sample consuming at least 4/7 of the following food groups (as a food or beverage) the previous day or night preceding the survey: grains, roots, tubers; pulses, nuts, seeds; dairy products; flesh foods; eggs; vitamin A-rich fruits and vegetables; other fruits and vegetables[27–29] | Children in the study sample |
| Minimum Dietary Diversity 8 (MDDC8) | Children aged 9-23 months in the study sample consuming at least 5/8 of the following food groups (as a food or beverage) the previous day or night preceding the survey: breastmilk; | Children aged 9-23 months in the study sample |

| | | |
|---|---|---|
| | grains, roots, tubers; pulses, nuts, seeds; dairy products; flesh foods; eggs; vitamin A-rich fruits and vegetables; other fruits and vegetables[27–29] | |
| Egg and/or Flesh Food Consumption | Children in the study sample consuming any eggs and/or animal-based flesh foods the previous day or night preceding the survey[29,30] | Children in the study sample |
| Fruit/Vegetable Consumption | Children in the study sample consuming any fruit and/or vegetables in the previous day or night preceding the survey[29,31] | Children in the study sample |
| Vitamin A-Rich Food Consumption | Children in the study sample consuming any of the following in the previous day or night preceding the survey: any vitamin A-rich fruits, tubers, or vegetables, dark green leafy vegetables, red palm oil foods, eggs, milk products, and/or animal organ meat[32] | Children in the study sample |
| Iron-Rich Food Consumption | Children in the study sample consuming any of the following in the previous day or night preceding the survey: dark green leafy vegetables, legumes, nuts, fish/seafood and/or any animal flesh foods[33] | Children in the study sample |
| Sugary Food Consumption | Children in the study sample consuming any sugary foods (chocolates/candies, cakes, pastries, etc.) the previous day or night preceding the survey[29,34] | Children in the study sample |
| ***Maternal- and Household-Related Key Indicators*** | | |
| **Indicator** | **Numerator** | **Denominator** |
| Household Decision-Making | Maternal: Households in the study sample where the mother was the decision-maker for income, healthcare, and purchases<br>Paternal: Households in the study sample where the father was the decision-maker for income, healthcare, and purchases<br>Joint: Households in the study sample where there was any combination of | Households in the study sample |

| | | |
|---|---|---|
| | mother and father decision-making for income, healthcare, and purchases | |
| Wealth/Asset Index[35] | Not Poor: Households in the study sample with wealth/asset indices falling into the middle, rich, or richest quintiles<br>Poor: Households in the study sample with wealth/asset indices falling in the poor or poorest quintile | Households in the study sample |
| Health- and Nutrition-Related Expenditure | Households in the study sample falling in the lower 25%, middle 50%, or upper 50% distribution when dividing household monthly spending on nutritious foods, medicine, and maternal and child health services by total household monthly expenditure and multiplying by 100 | Households in the study sample |
| Household Reduced Coping Strategy Index [36] | Households in the study sample scoring a reduced coping strategy index of <19 (IPC phase 1/2 (minimal/stressed)) or ≥19 (IPC phase 3/4/5 (crisis/emergency/famine)) | Households in the study sample |
| Household Crowding | Households in the study sample scoring a crowding index of at least 5, according to the number of individuals living in the household divided by the number of rooms in the household[37] | Households in the study sample |
| Household Food Consumption Score (FCS) [38] | Acceptable: Households in the study sample with a FCS score of 36+ (IPC Phase 1/2 (Minimal/Stressed))<br>Unacceptable: Households in the study sample with a FCS score <36 (IPC Phase 3/4/5 (Crisis/Emergency/Famine)) | Households in the study sample |
| Household Hunger Scale (HHS) [39] | Little-to-No Hunger: Households in the study sample with a HHS score of 0-1 (IPC Phase 1/2 (Minimal/Stressed))<br>Moderate-to-Severe Hunger: Households in the study sample with a HHS score of 2-6 (IPC Phase 3/4/5 (Crisis/Emergency/Famine)) | Households in the study sample |
| Use of Piped Water Source | Households in the study sample for which the main source of drinking | Households in the study sample |

| | | |
|---|---|---|
| | water for individuals in the household was from piped water or a public tap | |
| Use of Water Treatment | Households in the study sample for which the mother reports using at least one of the following water treatment methods prior to consumption/drinking of water: boil, chlorinate, keep under the sun, filter, use aquatab[40] | Households in the study sample |
| Household Open Defecation | Households in the study sample having no toilet facility and relying on bush/field for defecation[41] | Households in the study sample |
| Safe Disposal of Child's Stool | Households in the study sample for which the mother reports disposing of the child's stool in a latrine/toilet[42] | Households in the study sample |
| Safe Maternal Defecation Practices | Households in the study sample for which the mother reports using a latrine/toilet for defecation | Households in the study sample |
| Maternal Health-Related Knowledge | High: Households in the study sample where the mother had a score of 5-6 on the correct knowledge of the following 6 areas: exclusive breastfeeding, timing of breastfeeding initiation, timing of liquids initiation, timing of solid food initiation, handwashing moments, and water treatment methods<br>Low-to-Moderate: Households in the study sample where the mother had a score of 0-4 on the above areas of health-related knowledge | Households in the study sample |

**Text S4.** Construction of Different Indicators

**Wealth/Asset Index:** A wealth/asset index was calculated, incorporating multiple indicators representative of wealth in the study area.[35] From the questionnaire, ownership of livestock (camels, cattle, goats, donkeys, horses, poultry), household assets (electricity, radio, TV, telephone, computer, refrigerator, internet, air conditioning), and personal items (watch, mobile phone, bicycle, scooter, donkey cart, truck, canoe, tractor, ox plough) as well as household floor, roofing, and wall materials were considered.

Adapted binary variables were created for wealthier versus poorer floor, roofing, and wall materials. Ceramic, cement, wood, metal, and shingles roofs were considered wealthier for the purposes of this analysis, whereas having no roof or roofs constructed with leaves, grass, rustic mats, bamboo, wood planks, cardboard, cloth tent, or other materials were categorized as poorer.[10,43] Walls constructed of cement, stone, brick, cement blocks, wood planks/shingles, and covered adobe were considered wealthier in this analysis, whereas having no walls or walls constructed of cane, dirt, bamboo-mud, stone-mud, uncovered adobe, plywood, carboard, reused wood, or other materials were considered poorer.[10,43] Finally, polished, asphalt, tile, cement, and carpet floors were considered wealthier, whereas floors of earth, dung, wood planks, bamboo, or other materials were considered poorer.[10,43]

Only characteristics that were true of <95% or >5% of the study sample were considered (for example, if only 3% of the sample owned camels, camel ownership was excluded from construction of the wealth/asset index).[43] As the study sample was different for the CU5 and mothers' models at midline and the CU5 and mothers' models at endline, the wealth/asset index was calculated separately for each of these groups (calculated 4 times). Once variables to be considered were identified according to the described threshold, principal component analysis was performed with an acceptable minimum KMO threshold of 0.5.[44] Any variables falling below this threshold were excluded in a stepwise manner, with the variable with the lowest KMO value being excluded first until all variables were above the 0.5 KMO threshold. Quintiles were constructed, categorizing households into poorest, poor, middle, rich, and richest categories.[35,45] After examining the distribution of these quintiles in each study sample, a final binary variable of poor (combining the poor and poorest quintiles) and not poor (combining the middle, rich, and richest categories) was created.

The diagram below shows what variables were retained in each sample's wealth/asset index:

| Sample | Variables in Final Calculated Wealth/Asset Index |
|---|---|
| Midline CU5 Models | *Livestock Ownership:* Goats<br>*Agricultural Land Ownership*<br>*Household Assets:* Radio, Watch, Mobile Phone<br>*House Materials:* Roofing |
| Midline Mothers' Models | *Livestock Ownership:* Goats, Donkeys<br>*Agricultural Land Ownership*<br>*Household Assets:* Radio, Watch, Mobile Phone, Nonmobile Telephone<br>*House Materials:* Roofing, Walls |
| Endline CU5 Models | *Livestock Ownership:* Goats, Cattle<br>*Household Assets:* Radio<br>*House Materials:* Roofing, Walls, Flooring |
| Endline Mothers' Models | *Livestock Ownership:* Goats, Cattle<br>*Household Assets:* Radio<br>*Personal Assets:* Donkey Cart<br>*House Materials:* Roofing, Walls, Flooring |

**Health- and Nutrition-Related Expenditure:** A health- and nutrition-related expenditure variable was created by summing household monthly spending on nutritious foods, medicine, and maternal and child health (MCH) services (for example, antenatal care), dividing this value by total household monthly expenditure (including spending on nutritious foods, medicine, and MCH as well as other food, hygiene, transportation, fuel, water, electricity, rent, clothes, school, agriculture, social, debt, and savings), and multiplying by 100. Summary statistics for this variable were then examined, and a categorical variable was created to examine the lower 25%, middle 50%, and upper 25% categories.

**Household Decision-Making:** The questionnaire included separate questions about household decision-making on income, healthcare, and purchases. In order to avoid potential collinearity if including decision-making variables for each of these three components separately, a composite household decision-making variable was created by combining the answers to each of these areas, creating joint, maternal, and paternal household decision-making categories. A household was considered to have maternal household decision-making if the mother was listed as the decision-maker in all three areas of decision-making around income, healthcare, and purchases. Similarly, a household was considered to have paternal decision-making if the father was the reported decision-maker for income, healthcare, and purchases. Any other combination of decision-making around these three separate areas was considered joint household decision-making.

**Reduced Coping Strategy Index (rCSI):** The rCSI module contains questions around the use of the following coping mechanisms in the last seven days: relying on less preferred/less expensive foods, borrowing food, limiting portion sizes, restricting consumption by adults for children to eat, and reducing the number of meals per day.[36] The number of times each coping strategy was utilized in the last seven days was recorded, and a rCSI score was then calculated by adding up the number of days.[36] A binary variable testing rCSI scores of <19 and ≥19 was constructed, corresponding to phase 1/2 (minimal/stressed) versus phase 3/4/5 (crisis/emergency/famine), respectively, based on guidance from the Integrated Food Security Phase Classification (IPC) and as used by Save the Children.[46,47]

**Food Consumption Score (FCS):** The FCS module contains questions around days of consumption of the following food groups during the last seven days: cereals, legumes, vegetables, fruits, meat and fish, dairy, sugar, oil, and condiments.[38] Food groups were weighted and then summed to generate a FCS score, according to the following formula:

FCS = (days of cereals * 2) + (days of legumes * 3) + (days of vegetables) + (days of fruit) + (days of meat and fish * 4) + (days of dairy * 4) + (days of sugar * 0.5) + (days of oil * 0.5) + (days of condiments * 0)

FCS scores were then categorized into acceptable (>35) (corresponding to IPC Phase 1/2 (Minimal/Stressed)) or unacceptable (≤35) (corresponding to IPC Phase 3/4/5 (Crisis/Emergency/Famine)) levels of dietary diversity and food consumption.[38]

**Household Hunger Scale (HHS):** The HHS module was administered, asking respondents about the following experiences of household hunger in the 30 days preceding the survey: having no food available in the house, any household member going to sleep hungry due to a lack of available food in the house, and any household member going a full 24 hours without eating due to a lack of available food.[39] If respondents reported experiencing any of these events, they were asked to specify if the event occurred rarely, sometimes, or often in the past 30 days.[39] Responses of "never" experiencing the event were coded as 0, "rarely" or "sometimes" were coded as 1, and "often" was coded as 2.[39] These scores for each question were then summed to calculate a final HHS score.[39] These scores were then further categorized into the following groups: little-to-no hunger (scores of 0-1), moderate hunger (scores of 2-3), and severe hunger (scores of 4-6).[39] For the purposes of this analysis, the moderate and severe hunger categories were collapsed to create a binary indicator comparing moderate-severe household hunger (corresponding to IPC Phase 3/4/5 (Crisis/Emergency/Famine)) versus little-to-no household hunger (corresponding to IPC Phase 1/2 (Minimal/Stressed)).

**Household Crowding:** A household crowding index was calculated by dividing the number of individuals living in the household by the number of rooms in the household, as done elsewhere.[37] Responses greater than 3 for number of rooms in the household were re-coded as 3, for given the context, respondents were primarily living in tents with typically a maximum of 3 rooms (for example, a response of 22 rooms was recoded as 3). Crowding index values were then categorized into a binary variable of crowded (score $\geq 5$) versus not crowded (score $< 5$).

**Maternal Health-Related Knowledge:** A composite health-related knowledge variable was created, combining knowledge around exclusive breastfeeding, timing of breastfeeding initiation, timing of liquids initiation, timing of solid food initiation, handwashing moments, and water treatment methods. For each of these areas, a binary variable was created, coding correct knowledge as 1 and incorrect knowledge as 0. These six binary variables were then summed to arrive at a final composite knowledge score, ranging from 0-6, with 6 reflecting the best health-related knowledge. After examining the distribution of scores, a binary health-related knowledge variable was created to compare "high" knowledge (scores of 5-6) with "low-to-moderate" knowledge (scores of 0-4).

Exclusive breastfeeding knowledge was considered correct if a participant reported that infants should only receive breastmilk and no other liquids or solids for the first 6 months of life, and timing of breastfeeding initiation was correct if the participant identified that breastfeeding should be initiated within one hour of delivery.[48,49] Participants were considered to have correct knowledge on timing of liquids and solids introduction if they reported that introduction should occur when children are 6-8 months old.[49] Finally, correct knowledge of handwashing moments included if participants accurately identified at least 3/5 critical handwashing moments (before preparing food, before eating, before feeding children, after handling a child's stool/diaper, after using the latrine/toilet), and correct knowledge of water treatment methods included if participants identified at least one recommended method for treating water prior to consumption (boiling, chlorinating, keeping under the sun, filtering, using aquatab).[40,50]

**Minimum Dietary Diversity for Children (MDD-C):** MDD-C was calculated two ways- either including or excluding breastmilk as a food group - based on the age group of children and the relevance of breastmilk as an important dietary component.[27] If breastmilk was included, dietary diversity was considered to be met if infants consumed at least 5/8 food groups in the previous day.[27] If breastmilk was excluded, dietary diversity was met by consuming at least 4/7 food groups in the previous days.[27] While the present study included children aged 9-59 months old, the majority of children were aged 24-59 months (72.8% and 73.3% aged 24-59 months at midline and endline, respectively), with breastfeeding being less applicable for these older ages, as global recommendations encourage breastfeeding from birth to 2+ years, with complementary foods (starting at 6 months) comprising an increasingly larger share of nutritional requirements as children get older.[48] Therefore, to not underestimate dietary diversity, for the overall CU5 models and the stratified models of children aged 24-59 months, MDD-C was calculated without breastmilk as a food group (referred to in this analysis as MDDC7), as used elsewhere for children over 23 months.[51–53] For the stratified models of children 9-23 months old, MDD-C with breastmilk as a food group was calculated, as breastmilk is a critical food group for this age group (referred to as MDDC8 in this analysis), and this approach is recommended for children aged 6-23 months.[27]

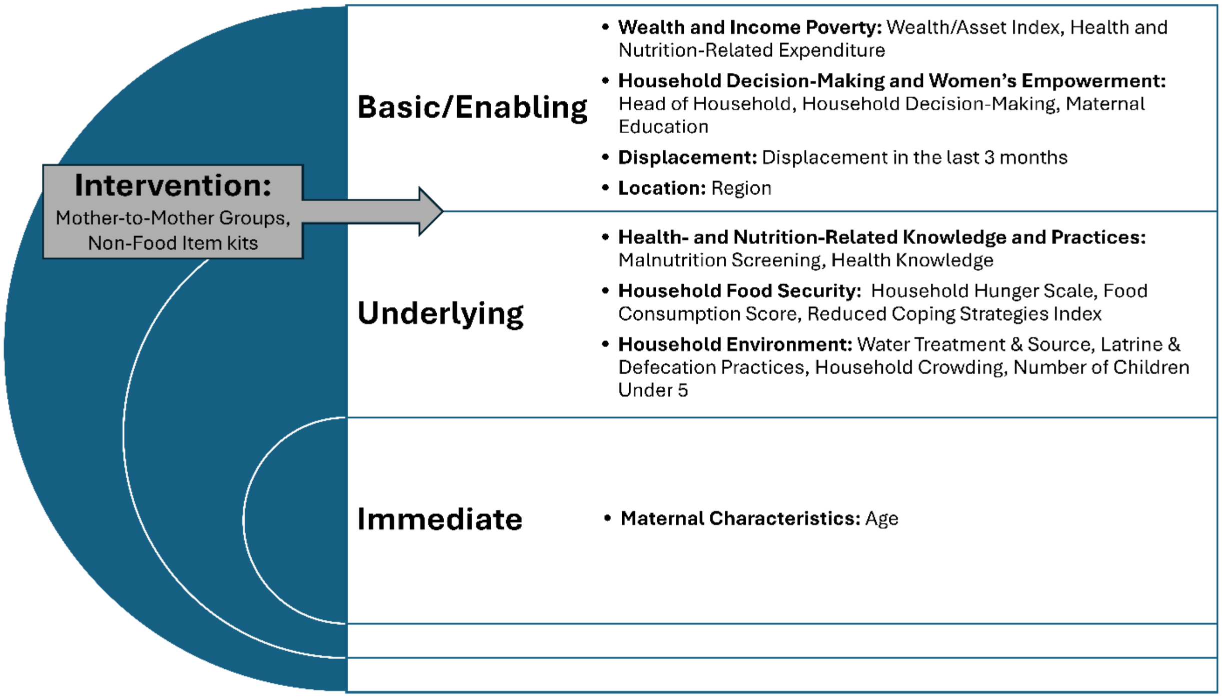


**Figure S1.** Mapping of Variables Tested in the Mothers' Models

**Text S5:** Defining the Study Sample

In the cRCT, data were collected on household conditions/factors, and anthropometric data on all CU5 (aged 6-59 months) in the household were collected; however, child characteristic questions around diet, disease, and vaccination status were only asked of the youngest child in each household.[8,9] Therefore, to be able to study these important factors, the child samples for this analysis focused only on the youngest child in each household aged 6-59 months. In the original data sets, there were 1263 youngest children-caregiver pairs at midline and 1145 youngest child-caregiver pairs at endline. Observations were removed if the reported caregiver was not the child's mother or if anthropometric data was missing. In both the child and mother datasets, mothers with reported ages above 65 were excluded from the analysis, as it was assumed that these women were likely the child's grandmother. In only the samples for the CU5 models, children were excluded if older than 59 months and if the child's WHZ was outside of the biologically plausible range of -5 to +5 SD. Additionally, children were excluded if the mother had given birth to a newborn since baseline to avoid any contamination around diet and disease questions concerning the youngest child in the household. In only the samples for the mothers' wasting analyses, mothers were excluded from the analysis if they had an extremely high (>60 cm) or low (8 cm) biologically implausible MUAC. This process resulted in final sample sizes of 956 children and 1066 mothers at midline and 833 children and 1023 mothers at endline.

**Text S6:** Modeling Strategy

A hierarchical model-building approach was used to assess the drivers of wasting, employing the basic/enabling, intervention, underlying, and immediate levels described in **Figure 1**.[54–56] During the first stage of analysis, crude, unadjusted bivariate relationships between the outcome of interest and each independent variable were examined, demonstrating the overall, crude relationship between each variable and the outcome (first column in each drivers results table). Next, major confounder-adjusted relationships were examined, adjusting for child sex and age, household region, and trial arm in the CU5 models and adjusting for household region and trial arm in the mothers' models (second column in each drivers results table). These major confounder-adjusted regression results depict the overall relationship between each variable and the outcome after adjusting for any potential confounding effects of child age, sex, household region, and trial arm.

From the major confounder-adjusted results, any independent variable that had an association with the outcome of interest with a p-value <0.20 was retained for the final hierarchical model-building approach, as used elsewhere.[57] A backwards-elimination model building approach was used and implemented in a hierarchical way at four levels (corresponding to Models 1-4 described below) according to our statistical framework (third column in each drivers results table).[56] Any independent variables from the basic/enabling level with a p-value <0.20 from the major confounder-adjusted association with the outcome of interest were entered into an initial regression model that adjusted for child age, sex, household region, and the trial arm in the child models and household region and trial arm in the mother models. Using a backwards elimination approach, variables with the highest p-values were removed one at a time until all remaining independent variables had a p-value <0.10. The coefficients, 95% confidence intervals, and p-values for these variables were reported (Model 1: basic/enabling drivers with p<0.10). Next, now ignoring the p-values of any retained basic/enabling variables, any intervention-related variables that had a p-value <0.20 from the adjusted associations were added to the model, again removing variables one at a time until all remaining intervention-related variables had a p-value <0.10. These coefficients, 95% confidence intervals, and p-values were then reported (Model 2: Model 1 + intervention variables with p<0.10). This process was then repeated with the underlying level variables (Model 3 = Model 2 + underlying variables with p<0.10) and then finally with the immediate level variables (Model 4 = Model 3 + immediate variables with p<0.10). Following the methods of Victora, this approach accounts for the hierarchical and interrelated nature of drivers (as characterized by the conceptual framework), allowing for the "true" association of a variable with the outcome to be reported without confounding by a more distal variable.[56] As a hypothetical example for illustrative purposes, a distal basic/enabling level variable, such as household wealth/assets, may influence child nutrition outcomes through more proximal variables at the underlying or immediate levels (for example, by influencing household hunger or child consumption of fruits/vegetables, respectively).[56] That said, by utilizing this hierarchical approach that progressively adjusts for drivers at more distal levels, the "true" association between child fruit/vegetable consumption and child wasting can be explored, having already accounted for more distal variables (such as wealth and household hunger).[56] That said, the results presented in the paper focus on the findings from the fully adjusted models using the hierarchical model building strategy, as these are believed to represent the most unbiased and unconfounded results for potential drivers by accounting for the interrelated hierarchical nature of the drivers of malnutrition.[56]

Throughout all analyses, a statistical significance level of $\alpha = 0.05$ is considered. Variables with a p-value above 0.05 and less than or equal to 0.10 were considered marginally statistically significant. For the purposes of this study, the cRCT trial arm was adjusted for in the analysis to control for any effect of the interventions. To not overfit models, the village cluster was not controlled for in this analysis.

For examining WHZ among CU5, an overall model with the entire sample of children was built with both the midline data and endline data (separately). Additional models were stratified by region, due to disproportionate wasting burdens by region, and by child age to assess drivers among children aged 9-23 months old versus 24-59 months old.

Regarding maternal wasting by MUAC, only overall models with all mothers were built at midline and endline. In contrast to child wasting, there was not a significant difference in the burden of wasting among mothers by region (chi-squared p-value = 0.182 at midline and 0.579 at endline).

For all final hierarchical multiple linear regression models for child WHZ (model 4 described above), Q-Q plots and Kernel Density Estimate plots of residuals were examined to assess normality of residuals, and residual-versus-fitted (RVF) plots and the Breusch-Pagan/Cook-Weisberg test were assessed to check for homoskedasticity. For any final hierarchical multiple linear regression models for child WHZ (model 4 described above) that had evidence of heteroskedasticity via the Breusch-Pagan/Cook-Weisberg test, the hierarchical four-stage model-building approach was repeated, using the "robust" option in STATA for robust standard errors. Additionally, histograms of difference in fits (DFITS) values were checked to identify any potential influential points. For all final hierarchical multivariable Poisson regression models (model 4 described above), Pearson and Deviance goodness-of-fit tests were performed. Collinearity among independent variables was checked in all final hierarchical adjusted models (model 4 described above) via assessing variance inflation factors (VIF). A conservative cutoff value of VIF $\geq 2.5$ was considered for potential collinearity.[58]

#### **Appendix S2.** Results Appendix

**Table S3.** Additional Child, Mother, and Household Characteristics*

| Level of Framework | Characteristic* | Midline | Endline |
|---|---|---|---|
| | *Child Minimum Dietary Diversity*** | n=956 | n=833 |
| | Acceptable | 370 (38.7%) | 347 (41.7%) |
| | Unacceptable | 586 (61.3%) | 486 (58.3%) |
| | *Child Egg and/or Flesh-Food Consumption* | n=956 | n=833 |
| | Consumed | 420 (43.9%) | 361 (43.3%) |
| | Not Consumed | 536 (56.1%) | 472 (56.7%) |
| | *Child Fruit/Vegetable Consumption* | n=956 | n=833 |
| | Consumed | 497 (52.0%) | 444 (53.3%) |
| | Not Consumed | 459 (48.0%) | 389 (46.7%) |
| ***Immediate*** | *Child Vitamin A-Rich Food Consumption* | n=956 | n=833 |
| | Consumed | 594 (62.1%) | 496 (59.5%) |
| | Not Consumed | 362 (37.9%) | 337 (40.5%) |
| | *Child Iron-Rich Food Consumption* | n=956 | n=833 |
| | Consumed | 587 (61.4%) | 501 (60.1%) |
| | Not Consumed | 369 (38.6%) | 332 (39.9%) |
| | *Child Sugary Food Consumption* | n=956 | n=833 |
| | Consumed | 556 (58.2%) | 466 (55.9%) |
| | Not Consumed | 400 (41.8%) | 367 (44.1%) |
| | *Maternal Pregnancy Status* | n=1066 | n=1022 |
| | Pregnant | 203 (19.0%) | 159 (15.6%) |
| | Not Pregnant | 863 (81.0%) | 863 (84.4%) |
| | *Child Tuberculosis Vaccination* | n=956 | n=833 |
| | Vaccinated | 442 (46.2%) | 534 (64.1%) |
| | Not Vaccinated | 514 (53.8%) | 299 (35.9%) |
| | *Maternal Health-Related Knowledge* | n=1066 | n=1023 |
| | High | 710 (66.6%) | 651 (63.6%) |
| | Low-Moderate | 356 (33.4%) | 372 (36.4%) |
| | *Household Number of CU5* | n=1066 | n=1023 |
| | 1 Child | 794 (74.5%) | 764 (74.7%) |
| | 2+ Children | 272 (25.5%) | 259 (25.3%) |
| | *Household Crowding* | n=1056 | -- |
| ***Underlying*** | Crowded | 423 (40.1%) | -- |
| | Not Crowded | 633 (59.9%) | -- |
| | *Household Reduced Coping Strategy Index* | n=1066 | n=1023 |
| | <19 (IPC Phase 1/2 (Minimal/Stressed)) | 826 (77.5%) | 726 (71.0%) |
| | ≥19 (IPC Phase 3/4/5 (Crisis/Emergency/Famine)) | 240 (22.5%) | 297 (29.0%) |
| | *Child Malnutrition Screening in Past 3 Months* | n=956 | n=833 |

| | | | |
|---|---|---|---|
| | Child Screened | 419 (43.8%) | 312 (37.5%) |
| | Child Not Screened | 537 (56.2%) | 521 (62.5%) |
| | *Mother Malnutrition Screening in Past 3 Months* | n=1066 | n=1023 |
| | Mother Screened | 649 (60.9%) | 694 (67.8%) |
| | Mother Not Screened | 417 (39.1%) | 329 (32.2%) |
| | *Household Use of Water Treatment* | n=1066 | n=1023 |
| | Yes | 837 (78.5%) | 880 (86.0%) |
| | No | 229 (21.5%) | 143 (14.0%) |
| | *Household Safe Disposal of Child's Stool* | n=1066 | n=1023 |
| | Open Area | 683 (64.1%) | 601 (58.8%) |
| | Latrine/Toilet | 383 (35.9%) | 422 (41.3%) |
| | *Safe Maternal Defecation Practices* | n=1066 | n=1023 |
| | Open Area | 325 (30.5%) | 429 (41.9%) |
| | Latrine/Toilet | 741 (69.5%) | 594 (58.1%) |
| ***Intervention*** | *Mother Attended Mother-to-Mother Support Groups* | n=1066 | n=1023 |
| | Attended | 516 (48.4%) | 737 (72.0%) |
| | Did Not Attend | 550 (51.6%) | 286 (28.0%) |
| | *Received Non-Food Item Kits* | n=1066 | n=1023 |
| | Yes | 193 (18.1%) | 380 (37.2%) |
| | No | 873 (81.9%) | 643 (62.9%) |
| ***Basic/Enabling*** | *Maternal Education* | -- | n=1022 |
| | Some Education | -- | 314 (30.7%) |
| | No Education | -- | 708 (69.3%) |
| | *Displacement in Last 3 Months* | -- | n=1023 |
| | Displaced | -- | 145 (14.2%) |
| | Not Displaced | -- | 878 (85.8%) |
| | *Head of Household* | -- | n=1022 |
| | Mother | -- | 575 (56.3%) |
| | Father or Other | -- | 447 (43.7%) |
| | *Health- and Nutrition-Related Expenditure* | n=1066 | n=1023 |
| | Lower 25% | 265 (24.9%) | 255 (24.9%) |
| | Middle 50% | 535 (50.2%) | 512 (50.1%) |
| | Upper 50% | 266 (25.0%) | 256 (25.0%) |

*Child characteristics are according to child samples at midline and endline. Mother, intervention, and household characteristics are according to mother samples at midline and endline. **MDDC7.

**Table S4.** Midline Overall CU5 Model: Drivers of Wasting (n=954)

| Domain/Indicator | Outcome: Wasting (Weight-for-Height Z-Score <-2 SD for children under five) | | |
|---|---|---|---|
| | Unadjusted Bivariate Poisson Regression Results | Major Confounder-Adjusted Poisson Regression Results** | Final Hierarchical Multivariable Poisson Regression Results*** |
| | RRR (95% CI)<br>p-value | RRR (95% CI)<br>p-value | RRR (95% CI)<br>p-value |
| **Basic/Enabling** | | | |
| ***Wealth and Income Poverty*** | | | |
| Wealth/Asset Index<br>(Ref: Not Poor) | 0.58 (0.40, 0.83)<br>0.003 | 1.07 (0.72, 1.59)<br>0.747 | -- |
| Health and Nutrition-Related Expenditure (Ref: Middle 50%) | | | |
| Lower 25% | 0.84 (0.53, 1.32)<br>0.449 | 1.00 (0.63, 1.60)<br>1.000 | -- |
| Upper 25% | 1.04 (0.68, 1.59)<br>0.848 | 0.84 (0.55, 1.28)<br>0.410 | -- |
| ***Household Decision-Making + Empowerment*** | | | |
| Decision-Making around Income, Purchases, & Healthcare (Ref: Joint) | | | |
| Maternal | 1.45 (0.93, 2.24)<br>0.099 | 1.11 (0.71, 1.73)<br>0.659 | -- |
| Paternal | 1.67 (1.10, 2.54)<br>0.016 | 1.06 (0.69, 1.63)<br>0.775 | -- |
| ***Region*** | | | |
| Hiran vs. Bay | 4.83 (2.86, 8.17)<br>0.000 | 4.74 (2.78, 8.10)<br>0.000 | 4.74 (2.78, 8.10)<br>0.000 |
| **Intervention** | | | |
| Attending M2M Groups<br>(Ref: Attended) | 1.35 (0.94, 1.94)<br>0.099 | 0.88 (0.60, 1.30)<br>0.530 | -- |
| NFI Kits<br>(Ref: Received) | *Not assessed in model due to 81.8% of the sample not receiving NFI kits* | | |
| **Underlying** | | | |
| ***Household Food Security*** | | | |
| Household Hunger Scale<br>(Ref: Little-to-no Hunger) | 1.57 (1.09, 2.27)<br>0.016 | 0.95 (0.65, 1.39)<br>0.789 | -- |
| Food Consumption Score<br>(Ref: Acceptable) | 1.22 (0.84, 1.76)<br>0.295 | 1.02 (0.70, 1.49)<br>0.910 | -- |
| Reduced Coping Strategy Index<br>(Ref: <19) | *Not assessed in model due to 77.2% of the sample having a rCSI <19* | | |
| ***Household Environment*** | | | |
| Household Crowding<br>(Ref: Not Crowded) | 2.21 (1.54, 3.17)<br>0.000 | 1.14 (0.76, 1.70)<br>0.527 | -- |
| Number of CU5<br>(Ref: 1 Child) | 0.83 (0.54, 1.25)<br>0.368 | 0.82 (0.52, 1.30)<br>0.399 | -- |
| Water Treatment<br>(Ref: Treats Water) | *Not assessed in models due to 80.3% of the sample treating water with an appropriate method* | | |
| Water Source<br>(Ref: Piped Water) | *Not assessed in model due to 77.6% of the sample not having household piped water* | | |
| Open Defecation<br>(Ref: Latrine/Toilet) | *Not assessed in models due to 76.7% of the sample not practicing open defecation* | | |
| Child Stool Disposal<br>(Ref: Latrine/Toilet) | 1.53 (1.03, 2.26)<br>0.036 | 1.48 (0.99, 2.22)<br>0.055 | 1.48 (0.99, 2.22)<br>0.055 |
| Maternal Defecation<br>(Ref: Latrine/Toilet) | 2.16 (1.52, 3.08)<br>0.000 | 1.45 (1.00, 2.09)<br>0.051 | -- |
| ***Health and Nutrition-Related Knowledge and Practices*** | | | |

| | | | |
|---|---|---|---|
| Tuberculosis Vaccination<br>(Ref: Vaccinated) | 2.00 (1.36, 2.94)<br>0.000 | 1.10 (0.72, 1.69)<br>0.646 | -- |
| Malnutrition Screening<br>(Ref: Child Screened) | 1.18 (0.82, 1.69)<br>0.373 | 1.18 (0.81, 1.70)<br>0.390 | -- |
| Health-Related Knowledge<br>(Ref: High) | 1.38 (0.96, 1.98)<br>0.082 | 1.14 (0.79, 1.64)<br>0.489 | -- |
| **Immediate** | | | |
| ***Diet*** | | | |
| Ever Breastfed<br>(Ref: Breastfed) | *Not assessed in models due to 81.5% of the sample having ever been breastfed* | | |
| Minimum Dietary Diversity<br>(Ref: Meets) | 1.87 (1.25, 2.82)<br>0.002 | 0.88 (0.56, 1.38)<br>0.570 | -- |
| *In the previous day, did not consume…* | | | |
| Fruit/Vegetables<br>(Ref: Consumed) | 2.09 (1.44, 3.03)<br>0.000 | 0.98 (0.64, 1.48)<br>0.907 | -- |
| Vitamin A Foods<br>(Ref: Consumed) | 1.72 (1.21, 2.45)<br>0.003 | 1.00 (0.69, 1.46)<br>0.980 | -- |
| Iron-Rich Foods<br>(Ref: Consumed) | 1.33 (0.93, 1.90)<br>0.116 | 0.97 (0.67, 1.39)<br>0.848 | -- |
| Eggs/Flesh Foods<br>(Ref: Consumed) | 1.27 (0.88, 1.82)<br>0.202 | 0.87 (0.59, 1.26)<br>0.459 | -- |
| *In the previous day, consumed…* | | | |
| Sugary Foods<br>(Ref: Not Consumes) | 0.83 (0.58, 1.19)<br>0.312 | 1.02 (0.71, 1.46)<br>0.914 | -- |
| ***Disease*** | | | |
| Illness in the Last 2 Weeks<br>(Ref: No Illness) | 1.26 (0.87, 1.81)<br>0.220 | 1.40 (0.97, 2.02)<br>0.072 | -- |
| ***Maternal Characteristics*** | | | |
| *Maternal MUAC (Ref: 24-29.5 cm)* | | | |
| Underweight (<24 cm) | 1.48 (0.96, 2.30)<br>0.077 | 1.55 (0.99, 2.41)<br>0.053 | 1.54 (0.99, 2.39)<br>0.057 |
| Overweight (>29.5 cm) | 0.66 (0.41, 1.05)<br>0.082 | 0.72 (0.45, 1.16)<br>0.177 | 0.77 (0.47, 1.24)<br>0.277 |
| *Maternal Age (Ref: 24-34 Years)* | | | |
| Younger (< 24) | 1.13 (0.66, 1.91)<br>0.658 | 1.31 (0.77, 2.23)<br>0.316 | -- |
| Older (35+) | 1.20 (0.82, 1.76)<br>0.358 | 1.16 (0.79, 1.72)<br>0.448 | -- |

*Note that final adjusted models may have a smaller sample size due to a few variables missing some observations. **Adjusting for child age, sex, household region, and trial arm. ***Adjusting for child age, sex, household region, and trial arm. Green indicates statistically significant at $p \leq 0.05$. Orange indicates marginally statistically significant from $0.05 < p \leq 0.10$. Pearson goodness-of-fit p-value for final fully adjusted model = 0.7986.

**Table S5.** Endline Overall CU5 Model: Drivers of Wasting (n=833)*

| **Domain/Indicator** | **Outcome: Wasting (Weight-for-Height Z-Score <-2 SD for children under five)** | | |
|---|---|---|---|
| | **Unadjusted Bivariate Poisson Regression Results** | **Major Confounder-Adjusted Poisson Regression Results**** | **Final Hierarchical Multivariable Poisson Regression Results***** |
| | RRR (95% CI) p-value | RRR (95% CI) p-value | RRR (95% CI) p-value |
| **Basic/Enabling** | | | |
| ***Wealth and Income Poverty*** | | | |
| Wealth/Asset Index (Ref: Not Poor) | 0.39 (0.25, 0.61) 0.000 | 0.75 (0.41, 1.37) 0.350 | -- |
| Health and Nutrition-Related Expenditure (Ref: Middle 50%) | | | |
| Lower 25% | 0.98 (0.63, 1.52) 0.914 | 1.08 (0.69, 1.70) 0.725 | -- |
| Upper 25% | 1.06 (0.69, 1.63) 0.797 | 0.85 (0.55, 1.32) 0.473 | -- |
| ***Household Decision-Making + Empowerment*** | | | |
| Head of Household (Ref: Mother) | 0.60 (0.41, 0.89) 0.012 | 0.75 (0.51, 1.12) 0.165 | -- |
| Decision-Making around Income, Purchases, & Healthcare (Ref: Joint) | | | |
| Maternal | 1.72 (1.15, 2.57) 0.009 | 1.41 (0.93, 2.12) 0.102 | -- |
| Paternal | 1.41 (0.84, 2.35) 0.192 | 1.14 (0.68, 1.91) 0.628 | -- |
| Maternal Education (Ref: Some Education) | 1.05 (0.71, 1.56) 0.789 | 1.19 (0.80, 1.76) 0.399 | -- |
| ***Displacement*** | | | |
| Displaced in Last 3 Months (Ref: Not Displaced) | *Not assessed due to 86.1% of the sample not being displaced* | | |
| ***Region*** | | | |
| Hiran vs. Bay | 2.82 (1.83, 4.35) 0.000 | 2.81 (1.80, 4.39) 0.000 | 2.81 (1.80, 4.39) 0.000 |
| **Intervention** | | | |
| Attending M2M Groups (Ref: Attended) | 0.78 (0.51, 1.19) 0.247 | 0.68 (0.44, 1.05) 0.085 | 0.68 (0.44, 1.05) 0.085 |
| NFI Kits (Ref: Received) | 0.61 (0.43, 0.88) 0.008 | 0.79 (0.55, 1.15) 0.226 | -- |
| **Underlying** | | | |
| ***Household Food Security*** | | | |
| Household Hunger Scale (Ref: Little-to-no Hunger) | 1.65 (1.14, 2.39) 0.008 | 1.20 (0.82, 1.78) 0.348 | -- |
| Food Consumption Score (Ref: Acceptable) | 1.23 (0.86, 1.75) 0.266 | 1.02 (0.71, 1.48) 0.897 | -- |
| Reduced Coping Strategy Index (Ref: <19) | 1.43 (0.99, 2.06) 0.059 | 1.27 (0.88, 1.85) 0.200 | -- |
| ***Household Environment*** | | | |
| Number of CU5 (Ref: 1 Child) | *Not assessed due to 75.2% of the sample having one child-under-5 in the home* | | |
| Water Treatment (Ref: Treats Water) | *Not assessed due to 88.1% of the sample treating water* | | |
| Water Source (Ref: Piped Water) | 1.14 (0.78, 1.67) 0.505 | 0.96 (0.65, 1.42) 0.828 | -- |
| Open Defecation | *Not assessed due to 76.1% of the sample not practicing open defecation* | | |

| | | | |
|---|---|---|---|
| (Ref: Latrine/Toilet) | | | |
| Child Stool Disposal<br>(Ref: Latrine/Toilet) | 1.69 (1.14, 2.50)<br>0.008 | 1.50 (1.01, 2.22)<br>0.045 | 1.53 (1.03, 2.27)<br>0.034 |
| Maternal Defecation<br>(Ref: Latrine/Toilet) | 1.99 (1.38, 2.87)<br>0.000 | 1.48 (1.01, 2.16)<br>0.043 | -- |
| ***Health and Nutrition-Related Knowledge and Practices*** | | | |
| Tuberculosis Vaccination<br>(Ref: Vaccinated) | 1.56 (1.09, 2.24)<br>0.015 | 1.06 (0.72, 1.55)<br>0.773 | -- |
| Malnutrition Screening<br>(Ref: Child Screened) | 1.58 (1.06, 2.36)<br>0.026 | 1.37 (0.91, 2.05)<br>0.130 | |
| Health-Related Knowledge<br>(Ref: High) | 1.25 (0.87, 1.80)<br>0.226 | 0.99 (0.68, 1.45)<br>0.970 | -- |
| **Immediate** | | | |
| ***Diet*** | | | |
| Ever Breastfed<br>(Ref: Breastfed) | *Not assessed due to 89.6% of the sample ever being breastfed* | | |
| Minimum Dietary Diversity<br>(Ref: Meets) | 1.54 (1.05, 2.26)<br>0.028 | 0.87 (0.55, 1.38)<br>0.550 | -- |
| *In the previous day, did not consume…* | | | |
| Fruit/Vegetables<br>(Ref: Consumed) | 1.30 (0.91, 1.87)<br>0.146 | 0.82 (0.55, 1.24)<br>0.357 | -- |
| Vitamin A Foods<br>(Ref: Consumed) | 1.52 (1.06, 2.18)<br>0.021 | 1.13 (0.77, 1.67)<br>0.531 | -- |
| Iron-Rich Foods<br>(Ref: Consumed) | 1.23 (0.86, 1.77)<br>0.251 | 0.93 (0.64, 1.36)<br>0.707 | -- |
| Eggs/Flesh Foods<br>(Ref: Consumed) | 1.86 (1.25, 2.75)<br>0.002 | 1.25 (0.80, 1.97)<br>0.332 | -- |
| *In the previous day, consumed…* | | | |
| Sugary Foods<br>(Ref: Not Consumed) | 0.56 (0.39, 0.81)<br>0.002 | 0.63 (0.44, 0.92)<br>0.017 | 0.67 (0.46, 0.98)<br>0.040 |
| ***Disease*** | | | |
| Illness in the Last 2 Weeks<br>(Ref: No Illness) | 1.23 (0.85, 1.80)<br>0.271 | 1.22 (0.84, 1.79)<br>0.293 | -- |
| ***Maternal Characteristics*** | | | |
| *Maternal MUAC (Ref: 24-29.5 cm)* | | | |
| Underweight (<24 cm) | 1.11 (0.71, 1.74)<br>0.634 | 1.13 (0.72, 1.77)<br>0.596 | -- |
| Overweight (>29.5 cm) | 0.90 (0.58, 1.39)<br>0.630 | 0.93 (0.60, 1.45)<br>0.743 | -- |
| *Maternal Age (Ref: 24-34 Years)* | | | |
| Younger (< 24) | 1.12 (0.65, 1.93)<br>0.674 | 1.31 (0.76, 2.26)<br>0.329 | -- |
| Older (35+) | 1.22 (0.83, 1.80)<br>0.320 | 1.20 (0.81, 1.79)<br>0.358 | -- |

*Note that final adjusted models may have a smaller sample size due to a few variables missing some observations. **Adjusting for child age, sex, household region, and trial arm. *** Adjusting for child age, sex, household region, and trial arm. Green indicates statistically significant at $p \leq 0.05$. Orange indicates marginally statistically significant from $0.05 < p \leq 0.10$. Pearson goodness-of-fit p-value for final fully adjusted model = 0.9974.

**Table S6.** Midline 9-23 Months Model: Drivers of Weight-for-Height Z-Score (n=260)*

| Domain/Indicator | Outcome: WHZ (Weight-for-Height Z-Score for children under five) | | |
|---|---|---|---|
| | **Unadjusted Bivariate Linear Regression Coefficient** | **Major Confounder-Adjusted Linear Regression Coefficient**** | **Final Hierarchical Multivariable Linear Regression Coefficient***** |
| | Coefficient estimate (95% CI) p-value | Coefficient estimate (95% CI) p-value | Coefficient estimate (95% CI) p-value |
| **Basic/Enabling** | | | |
| ***Wealth and Income Poverty*** | | | |
| Wealth/Asset Index (Ref: Not Poor) | -0.10 (-0.38, 0.18) 0.482 | -0.20 (-0.53, 0.13) 0.224 | -- |
| Health and Nutrition-Related Expenditure (Ref: Middle 50%) | | | |
| Lower 25% | -0.04 (-0.38, 0.30) 0.804 | 0.01 (-0.35, 0.36) 0.972 | -- |
| Upper 25% | 0.06 (-0.31, 0.42) 0.760 | 0.15 (-0.22, 0.51) 0.431 | -- |
| ***Household Decision-Making*** | | | |
| Decision-Making around Income, Purchases, & Healthcare (Ref: Joint) | | | |
| Maternal | 0.15 (-0.23, 0.53) 0.444 | 0.18 (-0.21, 0.56) 0.370 | -- |
| Paternal | -0.07 (-0.42, 0.29) 0.721 | 0.03 (-0.35, 0.41) 0.890 | -- |
| ***Region*** | | | |
| Hiran vs. Bay | -0.13 (-0.42, 0.15) 0.346 | -0.26 (-0.55, 0.02) 0.073 | -0.26 (-0.55, 0.02) 0.073 |
| **Intervention** | | | |
| Attending M2M Groups (Ref: Attended) | -0.01 (-0.29, 0.27) 0.953 | 0.23 (-0.07, 0.53) 0.139 | -- |
| NFI Kits (Ref: Received) | *Not assessed due to 82.3% of the sample not receiving NFI kits* | | |
| **Underlying** | | | |
| ***Household Food Security*** | | | |
| Household Hunger Scale (Ref: Little-to-no hunger) | *Not assessed due to 77.7% of the sample having little-to-no household hunger* | | |
| Food Consumption Score (Ref: Acceptable) | -0.22 (-0.51, 0.08) 0.147 | -0.14 (-0.44, 0.15) 0.344 | -- |
| Reduced Coping Strategy Index (Ref: <19) | 0.20 (-0.12, 0.52) 0.214 | 0.15 (-0.19, 0.49) 0.401 | -- |
| ***Household Environment*** | | | |
| Household Crowding (Ref: Not crowded) | 0.05 (-0.24, 0.34) 0.745 | 0.24 (-0.14, 0.61) 0.214 | -- |
| Number of CU5 (Ref: 1 Child) | 0.36 (0.08, 0.64) 0.011 | 0.36 (0.07, 0.64) 0.014 | 0.34 (0.06, 0.63) 0.019 |
| Water Treatment (Ref: Treated) | *Not assessed due to 81.5% of the sample practicing water treatment by an appropriate method* | | |
| Water Source (Ref: Piped) | 0.04 (-0.28, 0.36) 0.803 | 0.04 (-0.28, 0.36) 0.814 | -- |
| Open Defecation (Ref: Latrine/Toilet) | *Not assessed due to 82.3% of the sample not practicing open defecation* | | |
| Child Stool Disposal (Ref: Latrine) | -0.29 (-0.58, -0.00) 0.049 | -0.22 (-0.51, 0.07) 0.137 | -- |

| | | | |
|---|---|---|---|
| Maternal Defecation (Ref: Latrine) | -0.25 (-0.56, 0.07) 0.132 | -0.13 (-0.46, 0.20) 0.440 | -- |
| ***Health and Nutrition-Related Knowledge and Practices*** | | | |
| Tuberculosis Vaccination (Ref: Vaccinated) | -0.30 (-0.58, -0.02) 0.036 | -0.29 (-0.60, 0.02) 0.069 | -0.26 (-0.57, 0.05) 0.096 |
| Malnutrition Screening (Ref: Child Screened) | -0.04 (-0.32, 0.25) 0.806 | -0.05 (-0.34, 0.23) 0.707 | |
| Health and Nutrition Knowledge (Ref: High) | -0.02 (-0.32, 0.28) 0.890 | 0.04 (-0.25, 0.34) 0.772 | -- |
| **Immediate** | | | |
| ***Diet*** | | | |
| Ever Breastfed (Ref: Breastfed) | *Not assessed due to 86.5% of the sample having ever been breastfed* | | |
| Minimum Dietary Diversity (Ref: Met) | -0.10 (-0.41, 0.21) 0.516 | -0.05 (-0.39, 0.29) 0.771 | -- |
| *In the previous day, did not consume…* | | | |
| Fruit/Vegetables (Ref: Consumed) | -0.01 (-0.29, 0.27) 0.950 | 0.15 (-0.17, 0.48) 0.352 | -- |
| Vitamin A Foods (Ref: Consumed) | -0.07 (-0.35, 0.22) 0.638 | 0.07 (-0.25, 0.39) 0.661 | -- |
| Iron-Rich Foods (Ref: Consumed) | -0.01 (-0.29, 0.27) 0.950 | 0.02 (-0.28, 0.32) 0.884 | -- |
| Eggs/Flesh Foods (Ref: Consumed) | -0.11 (-0.39, 0.18) 0.469 | -0.05 (-0.35, 0.25) 0.745 | -- |
| *In the previous day, consumed…* | | | |
| Sugary Foods (Ref: Not Consumed) | 0.04 (-0.24, 0.33) 0.757 | 0.05 (-0.24, 0.33) 0.748 | -- |
| ***Disease*** | | | |
| Illness in the Last 2 Weeks (Ref: Not Ill) | -0.04 (-0.34, 0.26) 0.791 | -0.02 (-0.32, 0.27) 0.877 | -- |
| ***Maternal Characteristics*** | | | |
| *Maternal MUAC (Ref: 24-29.5 cm)* | | | |
| Underweight (<24 cm) | -0.35 (-0.75, 0.05) 0.083 | -0.31 (-0.71, 0.09) 0.124 | -- |
| Overweight (>29.5 cm) | 0.09 (-0.23, 0.41) 0.585 | 0.06 (-0.26, 0.37) 0.717 | -- |
| *Maternal Age (Ref: 24-34 Years)* | | | |
| Younger (<24) | -0.11 (-0.49, 0.26) 0.544 | -0.14 (-0.51, 0.23) 0.449 | -- |
| Older (35+) | 0.19 (-0.12, 0.51) 0.229 | 0.17 (-0.14, 0.49) 0.275 | -- |

*Note that final adjusted models may have a smaller sample size due to a few variables missing some observations. **Adjusting for child age, sex, household region, and trial arm. ***Adjusting for child age, sex, household region, and trial arm. Green indicates statistically significant at $p \leq 0.05$. Orange indicates marginally statistically significant from $0.05 < p \leq 0.10$.

**Table S7.** Midline 24-59 Months Model: Drivers of Weight-for-Height Z-Score (n=696)*

| **Domain/Indicator** | **Outcome: WHZ (Weight-for-Height Z-Score for children under five)** | | |
|---|---|---|---|
| | **Unadjusted Bivariate Linear Regression Coefficient** | **Major Confounder-Adjusted Linear Regression Coefficient**** | **Final Hierarchical Multivariable Linear Regression Coefficient***** |
| | Coefficient estimate (95% CI) p-value | Coefficient estimate (95% CI) p-value | Coefficient estimate (95% CI) p-value |
| **Basic/Enabling** | | | |
| ***Wealth and Income Poverty*** | | | |
| Wealth/Asset Index (Ref: Not Poor) | 0.49 (0.32, 0.65) 0.000 | 0.07 (-0.11, 0.25) 0.434 | -- |
| Health and Nutrition-Related Expenditure (Ref: Middle 50%): | | | |
| Lower 25% | 0.16 (-0.05, 0.37) 0.143 | 0.01 (-0.19, 0.21) 0.927 | -- |
| Upper 25% | -0.09 (-0.30, 0.11) 0.360 | 0.03 (-0.15, 0.22) 0.716 | -- |
| ***Household Decision-Making*** | | | |
| Decision-Making around Income, Purchases, & Healthcare (Ref: Joint) | | | |
| Maternal | -0.33 (-0.54, -0.12) 0.002 | -0.16 (-0.35, 0.03) 0.098 | -0.16 (-0.35, 0.03) 0.098 |
| Paternal | -0.38 (-0.59, -0.17) 0.000 | -0.09 (-0.28, 0.11) 0.386 | -0.09 (-0.28, 0.11) 0.386 |
| ***Region*** | | | |
| Hiran vs. Bay | -0.88 (-1.04, -0.73) 0.000 | -0.82 (-0.98, -0.66) 0.000 | -0.80 (-0.97, -0.64) 0.000 |
| **Intervention** | | | |
| Attending M2M Groups (Ref: Attended) | -0.20 (-0.37, -0.04) 0.018 | 0.02 (-0.15, 0.19) 0.820 | -- |
| NFI Kits (Ref: Received) | *Not assessed due to 81.6% of the sample not receiving NFI kits* | | |
| **Underlying** | | | |
| ***Household Food Security*** | | | |
| Household Hunger Scale (Ref: Little-to-no Hunger) | -0.41 (-0.60, -0.23) 0.000 | -0.00 (-0.19, 0.18) 0.962 | -- |
| Food Consumption Score (Ref: Acceptable) | -0.10 (-0.28, 0.09) 0.309 | -0.02 (-0.19, 0.15) 0.824 | -- |
| Reduced Coping Strategy Index (Ref: <19) | *Not assessed due to 78.3% of the sample having a rCSI <19* | | |
| ***Household Environment*** | | | |
| Household Crowding (Ref: Not Crowded) | -0.56 (-0.73, -0.39) 0.000 | -0.07 (-0.26, 0.13) 0.483 | -- |
| Number of CU5 (Ref: 1 Child) | *Not assessed due to 79.3% of the sample having one child-under-5 in the household* | | |
| Water Treatment (Ref: Treats Water) | *Not assessed due to 79.9% of the sample treating water with an appropriate method* | | |
| Water Source (Ref: Piped Water) | *Not assessed due to 78.7% of the sample not having household piped water* | | |
| Open Defecation (Ref: Latrine/Toilet) | -0.58 (-0.77, -0.39) 0.000 | -0.29 (-0.48, -0.10) 0.002 | -- |
| Child Stool Disposal (Ref: Latrine) | -0.17 (-0.35, 0.00) 0.054 | -0.20 (-0.36, -0.04) 0.016 | -- |

| | | | |
|---|---|---|---|
| Maternal Defecation<br>(Ref: Latrine) | -0.52 (-0.69, -0.34)<br>0.000 | -0.28 (-0.45, -0.11)<br>0.001 | -0.29 (-0.46, -0.11)<br>0.001 |
| ***Health and Nutrition-Related Knowledge and Practices*** | | | |
| Tuberculosis Vaccination<br>(Ref: Vaccinated) | -0.44 (-0.61, -0.27)<br>0.000 | 0.00 (-0.18, 0.18)<br>0.994 | -- |
| Malnutrition Screening<br>(Ref: Child Screened) | 0.02 (-0.15, 0.19)<br>0.847 | 0.01 (-0.15, 0.16)<br>0.949 | |
| Health-Related Knowledge<br>(Ref: High) | -0.17 (-0.35, 0.01)<br>0.065 | 0.04 (-0.13, 0.21)<br>0.674 | -- |
| **Immediate** | | | |
| ***Diet*** | | | |
| Ever Breastfed<br>(Ref: Breastfed) | *Not assessed due to 79.6% of the sample having ever breastfed* | | |
| Minimum Dietary Diversity<br>(Ref: Meets) | -0.59 (-0.75, -0.42)<br>0.000 | -0.13 (-0.31, 0.06)<br>0.183 | -- |
| *In the previous day, did not consume…* | | | |
| Fruit/Vegetables<br>(Ref: Consumed) | -0.58 (-0.75, -0.42)<br>0.000 | -0.06 (-0.25, 0.14)<br>0.554 | -- |
| Vitamin A Foods<br>(Ref: Consumed) | -0.44 (-0.61, -0.26)<br>0.000 | -0.02 (-0.20, 0.16)<br>0.823 | -- |
| Iron-Rich Foods<br>(Ref: Consumed) | -0.37 (-0.54, -0.19)<br>0.000 | -0.12 (-0.28, 0.05)<br>0.160 | -- |
| Eggs/Flesh Foods<br>(Ref: Consumed) | -0.31 (-0.48, -0.14)<br>0.000 | -0.06 (-0.22, 0.10)<br>0.451 | -- |
| *In the previous day, consumed…* | | | |
| Sugary Foods<br>(Ref: Not Consumed) | 0.17 (-0.00, 0.34)<br>0.056 | -0.03 (-0.19, 0.13)<br>0.711 | -- |
| ***Disease*** | | | |
| Illness in the Last 2 Weeks<br>(Ref: Not Ill) | -0.13 (-0.31, 0.05)<br>0.150 | -0.17 (-0.33, -0.00)<br>0.045 | -0.14 (-0.31, 0.03)<br>0.098 |
| ***Maternal Characteristics*** | | | |
| *Maternal MUAC (Ref: 24-29.5 cm)* | | | |
| Underweight (<24 cm) | -0.20 (-0.45, 0.05)<br>0.111 | -0.19 (-0.42, 0.03)<br>0.088 | -0.20 (-0.42, 0.02)<br>0.079 |
| Overweight (>29.5 cm) | 0.20 (0.00, 0.40)<br>0.046 | 0.18 (-0.00, 0.36)<br>0.052 | 0.14 (-0.04, 0.32)<br>0.123 |
| *Maternal Age (Ref: 24-34)* | | | |
| Younger (<24) | 0.00 (-0.26, 0.27)<br>0.986 | -0.02 (-0.26, 0.22)<br>0.872 | -- |
| Older (35+) | -0.04 (-0.22, 0.14)<br>0.654 | 0.01 (-0.15, 0.18)<br>0.860 | -- |

*Note that final adjusted models may have a smaller sample size due to a few variables missing some observations. **Adjusting for child age, sex, household region, and trial arm. ***Adjusting for child age, sex, household region, and trial arm. Green indicates statistically significant at $p \leq 0.05$. Orange indicates marginally statistically significant from $0.05 < p \leq 0.10$.

**Table S8.** Midline Hiran Region CU5 Model: Drivers of Weight-for-Height Z-Score (n=532)*

| Domain/Indicator | Outcome: WHZ (Weight-for-Height Z-Score for children under five) | | |
|---|---|---|---|
| | **Unadjusted Bivariate Linear Regression Coefficient** | **Major Confounder-Adjusted Linear Regression Coefficient**** | **Final Hierarchical Multivariable Linear Regression Coefficient***** |
| | Coefficient estimate (95% CI) p-value | Coefficient estimate (95% CI) p-value | Coefficient estimate (95% CI) p-value |
| **Basic/Enabling** | | | |
| ***Wealth and Income Poverty*** | | | |
| Wealth/Asset Index (Ref: Not Poor) | -0.05 (-0.26, 0.16) 0.611 | -0.06 (-0.26, 0.14) 0.547 | -- |
| Health and Nutrition-Related Expenditure (Ref: Middle 50%) | | | |
| Lower 25% | -0.01 (-0.29, 0.27) 0.947 | 0.06 (-0.20, 0.32) 0.665 | -- |
| Upper 25% | 0.10 (-0.11, 0.32) 0.344 | 0.11 (-0.09, 0.31) 0.285 | -- |
| ***Household Decision-Making*** | | | |
| Decision-Making around Income, Purchases, & Healthcare (Ref: Joint) | | | |
| Maternal | -0.38 (-0.62, -0.14) 0.002 | -0.27 (-0.50, -0.03) 0.024 | -0.27 (-0.50, -0.03) 0.024 |
| Paternal | -0.22 (-0.45, -0.00) 0.049 | -0.17 (-0.38, 0.04) 0.116 | -0.17 (-0.38, 0.04) 0.116 |
| **Intervention** | | | |
| Attending M2M Groups (Ref: Attended) | 0.03 (-0.17, 0.23) 0.791 | 0.10 (-0.10, 0.30) 0.326 | -- |
| NFI Kits (Ref: Received) | *Not assessed due to 79.1% of the sample not receiving NFI kits* | | |
| **Underlying** | | | |
| ***Household Food Security*** | | | |
| Household Hunger Scale (Ref: Little-to-no hunger) | -0.07 (-0.27, 0.12) 0.473 | -0.02 (-0.21, 0.16) 0.820 | -- |
| Food Consumption Score (Ref: Acceptable) | -0.21 (-0.41, -0.00) 0.045 | -0.19 (-0.38, 0.01) 0.060 | -- |
| Reduced Consumption Scale Index (Ref: <19) | *Not assessed due to 88.1% having a rCSI <19* | | |
| ***Household Environment*** | | | |
| Household Crowding (Ref: Not crowded) | -0.02 (-0.22, 0.19) 0.863 | -0.05 (-0.24, 0.15) 0.619 | -- |
| Number of CU5 (Ref: 1 Child) | 0.58 (0.39, 0.78) 0.000 | 0.59 (0.40, 0.79) 0.000 | 0.27 (0.06, 0.48) 0.011 |
| Water Treatment (Ref: Treated) | 0.06 (-0.15, 0.26) 0.575 | 0.04 (-0.15, 0.24) 0.665 | -- |
| Water Source (Ref: Piped Water) | *Not assessed due to 80.9% of the sample not having piped water* | | |
| Open Defecation (Ref: Latrine/Toilet) | -0.48 (-0.68, -0.28) 0.000 | -0.40 (-0.59, -0.20) 0.000 | -0.40 (-0.60, -0.20) 0.000 |
| Child Stool Disposal (Ref: Latrine) | -0.44 (-0.63, -0.25) 0.000 | -0.35 (-0.54, -0.16) 0.000 | -- |
| Maternal Defecation (Ref: Latrine) | -0.37 (-0.57, -0.18) 0.000 | -0.29 (-0.49, -0.10) 0.003 | -- |
| ***Health and Nutrition-Related Knowledge and Practices*** | | | |

| | | | |
|---|---|---|---|
| Tuberculosis Vaccination (Ref: Vaccinated) | *Not assessed due to 75.1% of the sample not being vaccinated against tuberculosis* | | |
| Malnutrition Screening (Ref: Child Screened) | 0.05 (-0.14, 0.25) 0.603 | 0.02 (-0.17, 0.21) 0.817 | -- |
| Health and Nutrition Knowledge (Ref: High) | 0.11 (-0.09, 0.31) 0.271 | 0.09 (-0.10, 0.28) 0.335 | -- |
| **Immediate** | | | |
| ***Diet*** | | | |
| Ever Breastfed (Ref: Breastfed) | -0.21 (-0.43, 0.01) 0.067 | -0.13 (-0.34, 0.08) 0.222 | |
| Minimum Dietary Diversity (Ref: Meets) | *Not assessed due to 83.1% of the sample not meeting dietary diversity* | | |
| *In the previous day, did not consume…* | | | |
| Fruit/Vegetables (Ref: Consumed) | -0.05 (-0.26, 0.17) 0.679 | -0.01 (-0.21, 0.20) 0.960 | -- |
| Vitamin A Foods (Ref: Consumed) | -0.02 (-0.22, 0.17) 0.838 | -0.04 (-0.22, 0.15) 0.703 | -- |
| Iron-Rich Foods (Ref: Consumed) | -0.09 (-0.28, 0.11) 0.384 | -0.15 (-0.33, 0.03) 0.103 | -- |
| Eggs/Flesh Foods (Ref: Consumed) | -0.13 (-0.34, 0.08) 0.218 | -0.18 (-0.38, 0.02) 0.072 | -- |
| *In the previous day, consumed…* | | | |
| Sugary Foods (Ref: Not Consumed) | 0.03 (-0.17, 0.22) 0.793 | 0.04 (-0.14, 0.22) 0.658 | -- |
| ***Disease*** | | | |
| Illness in the Last 2 Weeks (Ref: Not Ill) | -0.02 (-0.23, 0.19) 0.863 | -0.06 (-0.26, 0.14) 0.549 | -- |
| ***Maternal Characteristics*** | | | |
| *Maternal MUAC (Ref: 24-29.5 cm)* | | | |
| Underweight (<24 cm) | -0.32 (-0.60, -0.04) 0.023 | -0.36 (-0.62, -0.10) 0.006 | -0.39 (-0.65, -0.13) 0.004 |
| Overweight (>29.5 cm) | 0.28 (0.06, 0.51) 0.014 | 0.23 (0.01, 0.44) 0.039 | 0.12 (-0.09, 0.34) 0.258 |
| *Maternal Age (Ref: 24-34 Years)* | | | |
| Younger (<24) | -0.18 (-0.48, 0.12) 0.244 | -0.20 (-0.48, 0.08) 0.164 | -- |
| Older (35+) | -0.06 (-0.26, 0.15) 0.593 | 0.04 (-0.16, 0.24) 0.679 | -- |

*Note that final adjusted models may have a smaller sample size due to a few variables missing some observations. **Adjusting for child age, sex, household region, and trial arm. ***Adjusting for child age, sex, household region, and trial arm. Green indicates statistically significant at $p \leq 0.05$.

**Table S9.** Midline Bay Region CU5 Model: Drivers of Weight-for-Height Z-Score (n=401)*

| Domain/Indicator | Outcome: WHZ (Weight-for-Height Z-Score for children under five) | | |
|---|---|---|---|
| | **Unadjusted Bivariate Linear Regression Coefficient** | **Major Confounder-Adjusted Linear Regression Coefficient**** | **Final Hierarchical Multivariable Linear Regression Coefficient***** |
| | Coefficient estimate (95% CI) p-value | Coefficient estimate (95% CI) p-value | Coefficient estimate (95% CI) p-value |
| **Basic/Enabling** | | | |
| ***Wealth and Income Poverty*** | | | |
| Wealth/Asset Index (Ref: Not Poor) | *Not assessed due to 81.6% of the sample being in the poor category* | | |
| Health and Nutrition-Related Expenditure (Ref: Middle 50%) | | | |
| Lower 25% | -0.09 (-0.31, 0.13) 0.436 | -0.07 (-0.30, 0.16) 0.561 | -- |
| Upper 25% | -0.10 (-0.41, 0.21) 0.515 | -0.10 (-0.41, 0.21) 0.541 | -- |
| ***Household Decision-Making*** | | | |
| Decision-Making around Income, Purchases, & Healthcare (Ref: joint) | | | |
| Maternal | 0.17 (-0.08, 0.43) 0.188 | 0.18 (-0.08, 0.44) 0.180 | -- |
| Paternal | 0.18 (-0.16, 0.52) 0.295 | 0.23 (-0.11, 0.57) 0.187 | -- |
| **Intervention** | | | |
| Attending M2M Groups (Ref: Attended) | -0.00 (-0.21, 0.21) 0.994 | 0.04 (-0.18, 0.25) 0.747 | -- |
| NFI Kits (Ref: Received) | *Not assessed due to 85.5% of the sample not receiving NFI kits* | | |
| **Underlying** | | | |
| ***Household Food Security*** | | | |
| Household Hunger Scale (Ref: Little-to-no hunger) | *Not assessed due to 94.0% of the sample having little-to-no hunger* | | |
| Food Consumption Score (Ref: Acceptable) | 0.20 (-0.02, 0.43) 0.079 | 0.19 (-0.04, 0.41) 0.108 | -- |
| Reduced Consumption Score Index (Ref: <19) | -0.17 (-0.37, 0.04) 0.114 | -0.16 (-0.36, 0.05) 0.132 | -- |
| ***Household Environment*** | | | |
| Household Crowding (Ref: Not crowded) | *Not assessed due to 94.2% of the sample not having a crowded household* | | |
| Number of CU5 (Ref: 1 Child) | *Not assessed due to 82.5% of the sample having only 1 child under 5 in the home* | | |
| Water Treatment (Ref: Treats Water) | *Not assessed due to 98.3% of the sample treating water with an appropriate method* | | |
| Water Source (Ref: Piped) | 0.14 (-0.08, 0.37) 0.215 | 0.13 (-0.10, 0.36) 0.265 | -- |
| Open Defecation (Ref: Latrine/Toilet) | *Not assessed due to 90.8% of the sample not practicing open defecation* | | |
| Child Stool Disposal (Ref: Latrine) | -0.00 (-0.22, 0.21) 0.965 | 0.03 (-0.19, 0.25) 0.798 | -- |
| Maternal Defecation (Ref: Latrine) | *Not assessed due to 83% of the mothers using a latrine/toilet* | | |
| ***Health and Nutrition-Related Knowledge and Practices*** | | | |

| | | | |
|---|---|---|---|
| Tuberculosis Vaccination<br>(Ref: Vaccinated) | *Not assessed due to 75.8% of the sample being vaccinated against tuberculosis* | | |
| Malnutrition Screening<br>(Ref: Child Screened) | -0.01 (-0.22, 0.19)<br>0.886 | -0.02 (-0.23, 0.18)<br>0.810 | |
| Health-Related Knowledge<br>(Ref: High) | *Not assessed due to 78.6% of the sample having high health-related knowledge* | | |
| **Immediate** | | | |
| ***Diet*** | | | |
| Ever Breastfed<br>(Ref: Breastfed) | *Not assessed due to 91.3% of the sample having ever been breastfed* | | |
| Minimum Dietary Diversity<br>(Ref: Meets) | 0.02 (-0.19, 0.24)<br>0.841 | 0.02 (-0.20, 0.24)<br>0.867 | -- |
| *In the previous day, did not consume…* | | | |
| Fruit/Vegetables<br>(Ref: Consumed) | *Not assessed due to 86.8% consuming fruit/vegetables the previous day* | | |
| Vitamin A Foods<br>(Ref: Consumed) | *Not assessed due to 87.8% consuming Vitamin A foods the previous day* | | |
| Iron-Rich Foods<br>(Ref: Consumed) | *Not assessed due to 78.6% of the sample consuming Iron-rich foods the previous day* | | |
| Eggs/Flesh Foods<br>(Ref: Consumed) | 0.09 (-0.12, 0.29)<br>0.390 | 0.09 (-0.11, 0.30)<br>0.382 | -- |
| *In the previous day, consumed…* | | | |
| Sugary Foods<br>(Ref: Not Consumed) | -0.09 (-0.30, 0.13)<br>0.444 | -0.06 (-0.28, 0.16)<br>0.596 | -- |
| ***Disease*** | | | |
| Illness in the Last 2 Weeks<br>(Ref: Not Ill) | -0.27 (-0.47, -0.06)<br>0.013 | -0.26 (-0.47, -0.06)<br>0.013 | -0.26 (-0.47, -0.06)<br>0.013 |
| ***Maternal Characteristics*** | | | |
| *Maternal MUAC (Ref: 24-29.5 cm)* | | | |
| Underweight (<24 cm) | -0.08 (-0.37, 0.22)<br>0.610 | -0.06 (-0.35, 0.24)<br>0.694 | -- |
| Overweight (>29.5 cm) | 0.03 (-0.20, 0.26)<br>0.822 | 0.02 (-0.21, 0.25)<br>0.853 | -- |
| *Maternal Age (Ref: 24-34)* | | | |
| Younger (<24) | 0.07 (-0.21, 0.36)<br>0.609 | 0.08 (-0.21, 0.37)<br>0.582 | -- |
| Older (35+) | 0.06 (-0.16, 0.28)<br>0.571 | 0.07 (-0.15, 0.29)<br>0.532 | -- |

*Note that final adjusted models may have a smaller sample size due to a few variables missing some observations. **Adjusting for child age, sex, household region, and trial arm. ***Adjusting for child age, sex, household region, and trial arm. Green indicates statistically significant at $p \leq 0.05$.

**Table S10.** Endline 9-23 Month CU5 Model: Drivers of Weight-for-Height Z-Score (n=222)*

| **Domain/Indicator** | **Outcome: WHZ (Weight-for-Height Z-Score for children under five)** | | |
|---|---|---|---|
| | **Unadjusted Bivariate Linear Regression Coefficient** | **Major Confounder-Adjusted Linear Regression Coefficient**** | **Final Hierarchical Multivariable Linear Regression Coefficient***** |
| | Coefficient estimate (95% CI) p-value | Coefficient estimate (95% CI) p-value | Coefficient estimate (95% CI) p-value |
| **Basic/Enabling** | | | |
| ***Wealth and Income Poverty*** | | | |
| Wealth/Asset Index (Ref: Not Poor) | 0.08 (-0.23, 0.40) 0.600 | -0.00 (-0.45, 0.44) 0.991 | |
| Health and Nutrition-Related Expenditure (Ref: Middle 50%) | | | |
| Lower 25% | -0.09 (-0.47, 0.30) 0.664 | -0.17 (-0.56, 0.21) 0.378 | -- |
| Upper 25% | 0.15 (-0.23, 0.53) 0.428 | 0.22 (-0.16, 0.60) 0.248 | -- |
| ***Household Decision-Making + Empowerment*** | | | |
| Head of Household (Ref: Mother) | -0.13 (-0.44, 0.19) 0.429 | -0.15 (-0.47, 0.16) 0.345 | -- |
| Decision-Making around Income, Purchases, & Healthcare (Ref: Joint) | | | |
| Maternal | -0.04 (-0.44, 0.36) 0.845 | -0.05 (-0.47, 0.37) 0.809 | -- |
| Paternal | 0.42 (-0.07, 0.92) 0.092 | 0.41 (-0.09, 0.90) 0.104 | -- |
| Maternal Education (Ref: Some Education) | -0.54 (-0.87, -0.21) 0.001 | -0.59 (-0.92, -0.26) 0.001 | -0.59 (-0.93, -0.25) 0.001 |
| Displacement (Ref: Not Displaced) | *Not assessed due to 83.3% of the sample not being displaced in the last 3 months* | | |
| ***Region*** | | | |
| Hiran vs. Bay | -0.10 (-0.41, 0.22) 0.542 | -0.24 (-0.57, 0.08) 0.146 | -0.31 (-0.61, 0.00) 0.050 |
| **Intervention** | | | |
| Attending M2M Groups (Ref: Attended) | 0.11 (-0.25, 0.46) 0.553 | 0.24 (-0.12, 0.61) 0.193 | -- |
| NFI Kits (Ref: Received) | -0.12 (-0.44, 0.20) 0.455 | -0.05 (-0.38, 0.27) 0.749 | -- |
| **Underlying** | | | |
| ***Household Food Security*** | | | |
| Household Hunger Scale (Ref: Little-to-no Hunger) | 0.20 (-0.12, 0.51) 0.217 | 0.23 (-0.10, 0.56) 0.171 | -- |
| Food Consumption Score (Ref: Acceptable) | -0.21 (-0.52, 0.11) 0.195 | -0.24 (-0.57, 0.08) 0.142 | -- |
| Reduced Coping Strategy Index (Ref: <19) | -0.21 (-0.55, 0.14) 0.245 | -0.22 (-0.57, 0.12) 0.209 | -- |
| ***Household Environment*** | | | |
| Number of CU5 (Ref: 1 Child) | 0.12 (-0.19, 0.44) 0.450 | 0.13 (-0.20, 0.46) 0.440 | -- |
| Water Treatment (Ref: Treats Water) | *Not assessed due to 86.9% of the sample treating water* | | |
| Water Source (Ref: Piped Water) | 0.03 (-0.30, 0.36) 0.844 | 0.10 (-0.23, 0.44) 0.542 | -- |

| | | | |
|---|---|---|---|
| Open Defecation (Ref: Latrine/Toilet) | *Not assessed due to 79.7% of the sample not practicing open defecation* | | |
| Child Stool Disposal (Ref: Latrine/Toilet) | -0.21 (-0.52, 0.10) 0.188 | -0.23 (-0.54, 0.08) 0.149 | -- |
| Maternal Defecation (Ref: Latrine/Toilet) | -0.34 (-0.66, -0.02) 0.035 | -0.33 (-0.66, -0.00) 0.050 | -0.26 (-0.57, 0.05) 0.094 |
| ***Health and Nutrition-Related Knowledge and Practices*** | | | |
| Tuberculosis Vaccination (Ref: Vaccinated) | -0.28 (-0.62, 0.07) 0.115 | -0.21 (-0.58, 0.16) 0.271 | -- |
| Malnutrition Screening (Ref: Child Screened) | -0.13 (-0.45, 0.20) 0.445 | -0.16 (-0.49, 0.16) 0.320 | -- |
| Health-Related Knowledge (Ref: High) | 0.01 (-0.32, 0.33) 0.964 | 0.07 (-0.27, 0.41) 0.681 | -- |
| **Immediate** | | | |
| ***Diet*** | | | |
| Ever Breastfed (Ref: Breastfed) | *Not assessed due to 86.9% of the sample having been ever breastfed* | | |
| Minimum Dietary Diversity** (Ref: Meets) | -0.21 (-0.55, 0.12) 0.206 | -0.30 (-0.71, 0.10) 0.140 | -- |
| *In the previous day, did not consume…* | | | |
| Fruit/Vegetables (Ref: Consumed) | -0.15 (-0.46, 0.17) 0.355 | -0.18 (-0.60, 0.23) 0.385 | -- |
| Vitamin A Foods (Ref: Consumed) | -0.17 (-0.49, 0.14) 0.286 | -0.22 (-0.57, 0.14) 0.229 | -- |
| Iron-Rich Foods (Ref: Consumed) | 0.04 (-0.27, 0.36) 0.785 | 0.07 (-0.27, 0.41) 0.673 | -- |
| Eggs/Flesh Foods (Ref: Consumed) | -0.17 (-0.48, 0.14) 0.285 | -0.23 (-0.58, 0.13) 0.210 | -- |
| *In the previous day, consumed…* | | | |
| Sugary Foods (Ref: Not Consumed) | -0.11 (-0.42, 0.21) 0.502 | -0.05 (-0.38, 0.28) 0.766 | -- |
| ***Disease*** | | | |
| Illness in the Last 2 Weeks (Ref: Not Ill) | -0.25 (-0.59, 0.09) 0.156 | -0.25 (-0.59, 0.10) 0.157 | -- |
| ***Maternal Characteristics*** | | | |
| *Maternal MUAC (Ref: 24-29.5 cm)* | | | |
| Underweight (<24 cm) | 0.23 (-0.16, 0.62) 0.240 | 0.23 (-0.16, 0.63) 0.247 | 0.22 (-0.16, 0.61) 0.260 |
| Overweight (>29.5 cm) | 0.63 (0.26, 1.00) 0.001 | 0.57 (0.20, 0.94) 0.003 | 0.48 (0.10, 0.87) 0.015 |
| *Maternal Age (Ref: 24-34 Years)* | | | |
| Younger (<24) | -0.05 (-0.46, 0.37) 0.826 | -0.13 (-0.56, 0.29) 0.534 | -- |
| Older (35+) | -0.05 (-0.41, 0.31) 0.789 | -0.11 (-0.47, 0.25) 0.544 | -- |

*Note that final adjusted models may have a smaller sample size due to a few variables missing some observations. **Adjusting for child age, sex, household region, and trial arm. ***Adjusting for child age, sex, household region, and trial arm. Green indicates statistically significant at $p \leq 0.05$. Orange indicates marginally statistically significant from $0.05 < p \leq 0.10$.

**Table S11.** Endline 24-59 Month CU5 Model: Drivers of Weight-for-Height Z-Score (n=611)*

| **Domain/Indicator** | **Outcome: WHZ (Weight-for-Height Z-Score for children under five)** | | |
|---|---|---|---|
| | **Unadjusted Bivariate Linear Regression Coefficient** | **Major Confounder-Adjusted Linear Regression Coefficient**** | **Final Hierarchical Multivariable Linear Regression Coefficient***** |
| | Coefficient estimate (95% CI) p-value | Coefficient estimate (95% CI) p-value | Coefficient estimate (95% CI) p-value |
| **Basic/Enabling** | | | |
| ***Wealth and Income Poverty*** | | | |
| Wealth/Asset Index (Ref: Not Poor) | 0.46 (0.29, 0.63) 0.000 | -0.13 (-0.37, 0.10) 0.265 | |
| Health and Nutrition-Related Expenditure (Ref: Middle 50%) | | | |
| Lower 25% | -0.04 (-0.25, 0.17) 0.707 | -0.14 (-0.33, 0.06) 0.170 | -- |
| Upper 25% | -0.03 (-0.24, 0.19) 0.811 | 0.13 (-0.08, 0.33) 0.224 | -- |
| ***Household Decision-Making + Empowerment*** | | | |
| Head of Household (Ref: Mother) | 0.17 (-0.01, 0.34) 0.059 | -0.04 (-0.21, 0.13) 0.626 | -- |
| Decision-Making around Income, Purchases, & Healthcare (Ref: Joint) | | | |
| Maternal | -0.30 (-0.51, -0.10) 0.004 | -0.17 (-0.36, 0.03) 0.096 | -0.17 (-0.36, 0.03) 0.096 |
| Paternal | -0.37 (-0.62, -0.12) 0.003 | -0.20 (-0.44, 0.04) 0.095 | -0.20 (-0.44, 0.04) 0.095 |
| Maternal Education (Ref: Some Education) | 0.07 (-0.12, 0.26) 0.456 | -0.06 (-0.24, 0.11) 0.493 | -- |
| Displacement (Ref: Not Displaced) | *Not assessed due to 87.1% of the sample not being displaced in the last 3 months* | | |
| ***Region*** | | | |
| Hiran vs. Bay | -0.69 (-0.86, -0.53) 0.000 | -0.65 (-0.82, -0.48) 0.000 | -0.62 (-0.79, -0.45) 0.000 |
| **Intervention** | | | |
| Attending M2M Groups (Ref: Attended) | 0.14 (-0.05, 0.33) 0.162 | 0.14 (-0.05, 0.32) 0.148 | -- |
| NFI Kits (Ref: Received) | 0.26 (0.09, 0.43) 0.003 | 0.05 (-0.12, 0.22) 0.580 | -- |
| Underlying | | | |
| ***Household Food Security*** | | | |
| Household Hunger Scale (Ref: Little-to-no Hunger) | -0.28 (-0.45, -0.11) 0.001 | -0.01 (-0.18, 0.17) 0.936 | -- |
| Food Consumption Score (Ref: Acceptable) | -0.19 (-0.37, -0.02) 0.026 | -0.10 (-0.27, 0.06) 0.205 | -- |
| Reduced Coping Strategy Index (Ref: <19) | -0.18 (-0.37, 0.00) 0.054 | -0.08 (-0.25, 0.10) 0.377 | -- |
| ***Household Environment*** | | | |
| Number of CU5 (Ref: 1 Child) | *Not assessed due to 82.0% of the sample only having 1 CU5 in the household* | | |
| Water Treatment (Ref: Treats Water) | *Not assessed due to 88.5% of the sample treating water* | | |
| Water Source (Ref: Piped Water) | -0.13 (-0.31, 0.05) 0.157 | -0.02 (-0.19, 0.15) 0.819 | -- |

| | | | |
|---|---|---|---|
| Open Defecation<br>(Ref: Latrine/Toilet) | -0.04 (-0.24, 0.15)<br>0.657 | -0.00 (-0.19, 0.18)<br>0.963 | -- |
| Child Stool Disposal<br>(Ref: Latrine/Toilet) | -0.15 (-0.32, 0.03)<br>0.099 | -0.10 (-0.27, 0.06)<br>0.216 | -- |
| Maternal Defecation<br>(Ref: Latrine/Toilet) | -0.25 (-0.42, -0.08)<br>0.005 | -0.04 (-0.21, 0.12)<br>0.601 | -- |
| ***Health and Nutrition-Related Knowledge and Practices*** | | | |
| Tuberculosis Vaccination<br>(Ref: Vaccinated) | -0.15 (-0.33, 0.03)<br>0.095 | 0.14 (-0.04, 0.32)<br>0.121 | -- |
| Malnutrition Screening<br>(Ref: Child Screened) | -0.17 (-0.35, 0.00)<br>0.052 | -0.08 (-0.25, 0.08)<br>0.329 | -- |
| Health-Related Knowledge<br>(Ref: High) | -0.06 (-0.24, 0.12)<br>0.498 | 0.05 (-0.13, 0.22)<br>0.615 | -- |
| **Immediate** | | | |
| ***Diet*** | | | |
| Ever Breastfed<br>(Ref: Breastfed) | *Not assessed due to 90.5% of the sample having ever been breastfed* | | |
| Minimum Dietary Diversity<br>(Ref: Meets) | -0.35 (-0.52, -0.18)<br>0.000 | 0.05 (-0.15, 0.25)<br>0.640 | -- |
| *In the previous day, did not consume…* | | | |
| Fruit/Vegetables<br>(Ref: Consumed) | -0.31 (-0.48, -0.14)<br>0.000 | 0.03 (-0.17, 0.22)<br>0.793 | -- |
| Vitamin A Foods<br>(Ref: Consumed) | -0.28 (-0.45, -0.11)<br>0.002 | -0.04 (-0.22, 0.14)<br>0.691 | -- |
| Iron-Rich Foods<br>(Ref: Consumed) | -0.26 (-0.43, -0.08)<br>0.004 | -0.03 (-0.21, 0.14)<br>0.706 | -- |
| Eggs/Flesh Foods<br>(Ref: Consumed) | -0.48 (-0.65, -0.31)<br>0.000 | -0.22 (-0.41, -0.03)<br>0.022 | -0.24 (-0.43, -0.05)<br>0.014 |
| *In the previous day, consumed…* | | | |
| Sugary Foods<br>(Ref: Not Consumed) | 0.21 (0.04, 0.38)<br>0.016 | 0.06 (-0.11, 0.23)<br>0.487 | -- |
| ***Disease*** | | | |
| Illness in the Last 2 Weeks<br>(Ref: Not Ill) | -0.09 (-0.27, 0.10)<br>0.367 | -0.08 (-0.25, 0.10)<br>0.400 | -- |
| ***Maternal Characteristics*** | | | |
| *Maternal MUAC (Ref: 24-29.5 cm)* | | | |
| Underweight (<24 cm) | -0.10 (-0.32, 0.13)<br>0.404 | -0.09 (-0.30, 0.12)<br>0.422 | -- |
| Overweight (>29.5 cm) | 0.06 (-0.14, 0.26)<br>0.550 | 0.10 (-0.09, 0.29)<br>0.303 | -- |
| *Maternal Age (Ref: 24-34 Years)* | | | |
| Younger (<24) | 0.10 (-0.17, 0.37)<br>0.476 | 0.09 (-0.16, 0.35)<br>0.470 | -- |
| Older (35+) | 0.04 (-0.14, 0.22)<br>0.659 | 0.09 (-0.08, 0.26)<br>0.316 | -- |

*Note that final adjusted models may have a smaller sample size due to a few variables missing some observations. **Adjusting for child age, sex, household region, and trial arm. ***Adjusting for child age, sex, household region, and trial arm. Green indicates statistically significant at $p \leq 0.05$. Orange indicates marginally statistically significant from $0.05 < p \leq 0.10$.

**Table S12.** Endline Hiran Region CU5 Model: Drivers of Weight-for-Height Z-Score (n=468)*

| **Domain/Indicator** | **Outcome: WHZ (Weight-for-Height Z-Score for children under five)** | | |
|---|---|---|---|
| | **Unadjusted Bivariate Linear Regression Coefficient** | **Major Confounder-Adjusted Linear Regression Coefficient**** | **Final Hierarchical Multivariable Linear Regression Coefficient***** |
| | Coefficient estimate (95% CI) p-value | Coefficient estimate (95% CI) p-value | Coefficient estimate (95% CI) p-value |
| **Basic/Enabling** | | | |
| ***Wealth and Income Poverty*** | | | |
| Wealth/Asset Index (Ref: Not Poor) | *Not assessed due to 90.8% of the sample being in the "not poor" category* | | |
| Health and Nutrition-Related Expenditure (Ref: Middle 50%) | | | |
| Lower 25% | -0.16 (-0.43, 0.11) 0.248 | -0.16 (-0.41, 0.10) 0.224 | -- |
| Upper 25% | 0.07 (-0.16, 0.30) 0.552 | 0.15 (-0.07, 0.38) 0.180 | -- |
| ***Household Decision-Making + Empowerment*** | | | |
| Head of Household (Ref: Mother) | 0.13 (-0.08, 0.35) 0.224 | 0.12 (-0.09, 0.33) 0.257 | -- |
| Decision-Making around Income, Purchases, & Healthcare (Ref: Joint) | | | |
| Maternal | -0.07 (-0.31, 0.17) 0.561 | -0.06 (-0.28, 0.17) 0.621 | -- |
| Paternal | 0.04 (-0.23, 0.31) 0.778 | 0.01 (-0.25, 0.27) 0.945 | -- |
| Maternal Education (Ref: Some Education) | -0.13 (-0.33, 0.08) 0.240 | -0.10 (-0.30, 0.10) 0.312 | -- |
| Displacement (Ref: Not Displaced) | *Not assessed due to 98.7% of the sample not being displaced in the last 3 months* | | |
| **Intervention** | | | |
| Attending M2M Groups (Ref: Attended) | 0.05 (-0.18, 0.28) 0.666 | 0.19 (-0.04, 0.41) 0.105 | -- |
| NFI Kits (Ref: Receiving NFI Kit) | 0.16 (-0.04, 0.36) 0.120 | 0.21 (0.01, 0.40) 0.035 | 0.21 (0.01, 0.40) 0.035 |
| **Underlying** | | | |
| ***Household Food Security*** | | | |
| Household Hunger Scale (Ref: Little-to-no Hunger) | -0.04 (-0.25, 0.17) 0.683 | -0.01 (-0.21, 0.19) 0.943 | -- |
| Food Consumption Score (Ref: Acceptable) | -0.07 (-0.27, 0.14) 0.516 | -0.04 (-0.23, 0.16) 0.720 | -- |
| Reduced Coping Strategy Index (Ref: <19) | -0.16 (-0.38, 0.05) 0.134 | -0.06 (-0.27, 0.14) 0.534 | -- |
| ***Household Environment*** | | | |
| Number of CU5 (Ref: 1 Child) | 0.36 (0.15, 0.58) 0.001 | 0.08 (-0.15, 0.30) 0.493 | -- |
| Water Treatment (Ref: Treats Water) | *Not assessed due to 84.8% of the sample treating water* | | |
| Water Source (Ref: Piped Water) | -0.07 (-0.29, 0.15) 0.536 | -0.08 (-0.30, 0.13) 0.454 | -- |
| Open Defecation (Ref: Latrine/Toilet) | -0.15 (-0.38, 0.08) 0.190 | -0.03 (-0.26, 0.19) 0.783 | -- |
| Child Stool Disposal | -0.15 (-0.36, 0.05) | -0.09 (-0.29, 0.11) | -- |

| | | | |
|---|---|---|---|
| (Ref: Latrine/Toilet) | 0.143 | 0.385 | |
| Maternal Defecation<br>(Ref: Latrine/Toilet) | -0.28 (-0.48, -0.08)<br>0.007 | -0.19 (-0.38, 0.00)<br>0.055 | -0.17 (-0.36, 0.01)<br>0.065 |
| ***Health and Nutrition-Related Knowledge and Practices*** | | | |
| Tuberculosis Vaccination<br>(Ref: Vaccinated) | 0.04 (-0.16, 0.24)<br>0.714 | 0.13 (-0.07, 0.32)<br>0.196 | -- |
| Malnutrition Screening<br>(Ref: Child Screened) | -0.12 (-0.34, 0.09)<br>0.253 | -0.08 (-0.28, 0.13)<br>0.453 | |
| Health-Related Knowledge<br>(Ref: High) | -0.06 (-0.26, 0.15)<br>0.596 | -0.02 (-0.22, 0.18)<br>0.871 | -- |
| **Immediate** | | | |
| ***Diet*** | | | |
| Ever Breastfed<br>(Ref: Breastfed) | *Not assessed due to 90.6% of the sample ever being breastfed* | | |
| Minimum Dietary Diversity<br>(Ref: Meets) | *Not assessed due to 83.8% of the sample not being minimum dietary diversity* | | |
| *In the previous day, did not consume…* | | | |
| Fruit/Vegetables<br>(Ref: Consumed) | 0.23 (0.01, 0.46)<br>0.039 | 0.12 (-0.10, 0.34)<br>0.282 | -- |
| Vitamin A Foods<br>(Ref: Consumed) | 0.04 (-0.16, 0.25)<br>0.694 | -0.01 (-0.21, 0.18)<br>0.900 | -- |
| Iron-Rich Foods<br>(Ref: Consumed) | 0.14 (-0.07, 0.34)<br>0.184 | 0.12 (-0.07, 0.31)<br>0.224 | -- |
| Eggs/Flesh Foods<br>(Ref: Consumed) | *Not assessed due to 78.6% of the sample not eating egg and/or flesh foods the previous day* | | |
| *In the previous day, consumed…* | | | |
| Sugary Foods<br>(Ref: Not Consumed) | -0.04 (-0.24, 0.16)<br>0.712 | 0.01 (-0.18, 0.20)<br>0.919 | -- |
| ***Disease*** | | | |
| Illness in the Last 2 Weeks<br>(Ref: Not Ill) | -0.04 (-0.26, 0.18)<br>0.702 | -0.04 (-0.25, 0.17)<br>0.685 | -- |
| ***Maternal Characteristics*** | | | |
| *Maternal MUAC (Ref: 24-29.5 cm)* | | | |
| Underweight (<24 cm) | -0.12 (-0.38, 0.15)<br>0.383 | -0.10 (-0.35, 0.15)<br>0.451 | -- |
| Overweight (>29.5 cm) | 0.17 (-0.07, 0.40)<br>0.171 | 0.19 (-0.04, 0.42)<br>0.111 | -- |
| *Maternal Age (Ref: 24-34 Years)* | | | |
| Younger (<24) | 0.01 (-0.31, 0.33)<br>0.934 | -0.01 (-0.32, 0.29)<br>0.935 | -- |
| Older (35+) | -0.07 (-0.29, 0.14)<br>0.510 | 0.03 (-0.18, 0.24)<br>0.792 | -- |

*Note that final adjusted models may have a smaller sample size due to a few variables missing some observations. **Adjusting for child age, sex, household region, and trial arm. ***Adjusting for child age, sex, household region, and trial arm. Green indicates statistically significant at $p \leq 0.05$. Orange indicates marginally statistically significant from $0.05 < p \leq 0.10$.

**Table S13.** Endline Bay CU5 Model: Drivers of Weight-for-Height Z-Score (n=364)*

| Domain/Indicator | Outcome: WHZ (Weight-for-Height Z-Score for children under five) | | |
|---|---|---|---|
| | **Unadjusted Bivariate Linear Regression Coefficient** | **Major Confounder-Adjusted Linear Regression Coefficient**** | **Final Hierarchical Multivariable Linear Regression Coefficient***** |
| | Coefficient estimate (95% CI) p-value | Coefficient estimate (95% CI) p-value | Coefficient estimate (95% CI) p-value |
| **Basic/Enabling** | | | |
| ***Wealth and Income Poverty*** | | | |
| Wealth/Asset Index (Ref: Not Poor) | *Not assessed due to 79.7% of the sample being in the "poor" category* | | |
| Health and Nutrition-Related Expenditure (Ref: Middle 50%) | | | |
| Lower 25% | -0.08 (-0.33, 0.16) 0.517 | -0.06 (-0.30, 0.19) 0.640 | -- |
| Upper 25% | 0.18 (-0.12, 0.48) 0.247 | 0.16 (-0.14, 0.46) 0.290 | -- |
| ***Household Decision-Making + Empowerment*** | | | |
| Head of Household (Ref: Mother) | -0.19 (-0.41, 0.02) 0.080 | -0.20 (-0.43, 0.02) 0.070 | -0.31 (-0.56, -0.06) 0.014 |
| Decision-Making around Income, Purchases, & Healthcare (Ref: Joint) | | | |
| Maternal | -0.27 (-0.56, 0.02) 0.069 | -0.24 (-0.53, 0.05) 0.110 | -0.41 (-0.74, -0.09) 0.012 |
| Paternal | -0.28 (-0.68, 0.12) 0.172 | -0.35 (-0.76, 0.06) 0.092 | -0.26 (-0.67, 0.16) 0.222 |
| Maternal Education (Ref: Some Education) | *Not assessed due to 76.1% of the sample's mothers not having any education* | | |
| ***Displacement*** | | | |
| Displaced in Last 3 Months (Ref: Not Displaced) | 0.05 (-0.18, 0.29) 0.666 | 0.01 (-0.24, 0.26) 0.926 | -- |
| **Intervention** | | | |
| Attending M2M Groups (Ref: Attended) | 0.18 (-0.05, 0.42) 0.129 | 0.15 (-0.10, 0.40) 0.252 | -- |
| NFI Kits (Ref: Received) | *Not assessed due to 78.6% of the sample not receiving NFI kits* | | |
| **Underlying** | | | |
| ***Household Food Security*** | | | |
| Household Hunger Scale (Ref: Little-to-no Hunger) | 0.13 (-0.11, 0.36) 0.286 | 0.15 (-0.09, 0.38) 0.217 | -- |
| Food Consumption Score (Ref: Acceptable) | -0.17 (-0.39, 0.05) 0.131 | -0.14 (-0.37, 0.09) 0.221 | -- |
| Reduced Coping Strategy Index (Ref: <19 | -0.15 (-0.39, 0.10) 0.245 | -0.11 (-0.36, 0.14) 0.380 | |
| ***Household Environment*** | | | |
| Number of CU5 (Ref: 1 Child) | *Not assessed due to 84.7% of the sample having 1 child-under-5 in the household* | | |
| Water Treatment (Ref: Treats Water) | *Not assessed due to 92.3% of the sample treating water* | | |
| Water Source (Ref: Piped Water) | 0.07 (-0.15, 0.28) 0.555 | 0.05 (-0.18, 0.28) 0.686 | -- |
| Open Defecation (Ref: Latrine/Toilet) | *Not assessed due to 78.6% of the sample not practicing open defecation* | | |

| | | | |
|---|---|---|---|
| Child Stool Disposal<br>(Ref: Latrine/Toilet) | -0.11 (-0.32, 0.11)<br>0.336 | -0.08 (-0.31, 0.14)<br>0.473 | -- |
| Maternal Defecation<br>(Ref: Latrine/Toilet) | 0.04 (-0.20, 0.28)<br>0.728 | 0.05 (-0.19, 0.29)<br>0.698 | -- |
| ***Health and Nutrition-Related Knowledge and Practices*** | | | |
| Tuberculosis Vaccination<br>(Ref: Vaccinated) | *Not assessed due to 83.3% of the sample being vaccinated against tuberculosis* | | |
| Malnutrition Screening<br>(Ref: Child Screened) | -0.08 (-0.30, 0.14)<br>0.456 | -0.09 (-0.31, 0.13)<br>0.404 | |
| Health-Related Knowledge<br>(Ref: High) | 0.21 (-0.03, 0.45)<br>0.089 | 0.18 (-0.08, 0.44)<br>0.164 | -- |
| **Immediate** | | | |
| ***Diet*** | | | |
| Ever Breastfed<br>(Ref: Breastfed) | *Not assessed due to 88.2% of the sample having ever been breastfed* | | |
| Minimum Dietary Diversity<br>(Ref: Meets) | -0.08 (-0.33, 0.16)<br>0.506 | -0.09 (-0.34, 0.16)<br>0.480 | -- |
| *In the previous day, did not consume…* | | | |
| Fruit/Vegetables<br>(Ref: Consumed) | *Not assessed due to 84.7% of the sampling eating fruits/vegetables the previous day* | | |
| Vitamin A Foods<br>(Ref: Consumed) | *Not assessed due to 82.2% of the sample consuming Vitamin A-rich foods the previous day* | | |
| Iron-Rich Foods<br>(Ref: Consumed) | *Not assessed due to 79.2% of the sample consuming Iron-rich foods the previous day* | | |
| Eggs/Flesh Foods<br>(Ref: Consumed) | -0.10 (-0.34, 0.14)<br>0.429 | -0.10 (-0.35, 0.15)<br>0.443 | -- |
| *In the previous day, consumed…* | | | |
| Sugary Foods<br>(Ref: Not Consumed) | 0.05 (-0.19, 0.28)<br>0.698 | 0.02 (-0.22, 0.26)<br>0.845 | -- |
| ***Disease*** | | | |
| Illness in the Last 2 Weeks<br>(Ref: Not Ill) | -0.23 (-0.46, 0.01)<br>0.060 | -0.22 (-0.46, 0.01)<br>0.063 | -- |
| ***Maternal Characteristics*** | | | |
| *Maternal MUAC (Ref: 24-29.5 cm)* | | | |
| Underweight (<24 cm) | 0.14 (-0.14, 0.42)<br>0.324 | 0.12 (-0.16, 0.40)<br>0.396 | 0.12 (-0.16, 0.39)<br>0.409 |
| Overweight (>29.5 cm) | 0.25 (-0.01, 0.50)<br>0.059 | 0.24 (-0.02, 0.49)<br>0.069 | 0.28 (0.03, 0.54)<br>0.030 |
| *Maternal Age (Ref: 24-34 Years)* | | | |
| Younger (<24) | 0.04 (-0.26, 0.35)<br>0.775 | 0.04 (-0.27, 0.35)<br>0.797 | -- |
| Older (35+) | 0.06 (-0.18, 0.30)<br>0.612 | 0.08 (-0.16, 0.32)<br>0.526 | -- |

*Note that final adjusted models may have a smaller sample size due to a few variables missing some observations. **Adjusting for child age, sex, household region, and trial arm. ***Adjusting for child age, sex, household region, and trial arm. Green indicates statistically significant at $p \leq 0.05$.

**Drivers of Wasting (MUAC<23cm)**

<u>*Basic/Enabling*</u>
- Decision-Making$^{M}$

<u>*Intervention*</u>
- M2M Groups$^{E}$

<u>*Underlying*</u>
- Household Hunger Scale$^{M}$
- Food Consumption Score$^{M}$
- Child Stool Disposal$^{M}$
- Open Defecation$^{E}$

<u>*Immediate*</u>
-Age$^{M}$

**Figure S2.** Drivers of Mothers' Wasting

*"M" superscript indicates the variable was a statistically significant driver in the given model at midline, and "E" superscript indicates the variable was a driver in the given model at endline. Note that this image depicts statistically significant drivers from the final adjusted models at the α = 0.05 level; additional drivers may have been marginally statistically significant or statistically significant in the unadjusted or basic confounder-adjusted models.*

**Table S14.** Midline Overall Maternal MUAC Model: Drivers of Wasting (n=1066)*

| Domain/Indicator | Outcome: Wasting (Mid-upper arm circumference <23 cm of mothers) | | |
|---|---|---|---|
| | **Unadjusted Bivariate Poisson Regression Coefficient** | **Major Confounder-Adjusted Poisson Regression Coefficient**** | **Final Hierarchical Multivariable Poisson Regression Coefficient***** |
| | Relative Risk Ratio (95% CI) p-value | Relative Risk Ratio (95% CI) p-value | Relative Risk Ratio (95% CI) p-value |
| **Basic/Enabling** | | | |
| ***Wealth and Income Poverty*** | | | |
| Wealth/Asset Index (Ref: Not Poor) | 1.16 (0.76, 1.77) 0.486 | 1.42 (0.89, 2.25) 0.142 | -- |
| Health- and Nutrition-Related Expenditure (Ref: Middle 50%) | | | |
| Lower 25% | 1.42 (0.86, 2.34) 0.172 | 1.49 (0.88, 2.51) 0.136 | 1.59 (0.94, 2.68) 0.085 |
| Upper 25% | 1.30 (0.78, 2.18) 0.310 | 1.23 (0.73, 2.06) 0.441 | 1.20 (0.71, 2.02) 0.495 |
| ***Household Decision-Making*** | | | |
| Decision-Making around Income, Purchases, & Healthcare (Ref: Joint) | | | |
| Maternal | 0.54 (0.29, 1.04) 0.066 | 0.52 (0.27, 0.99) 0.047 | 0.49 (0.26, 0.95) 0.034 |
| Paternal | 1.00 (0.61, 1.66) 0.988 | 0.90 (0.53, 1.52) 0.689 | 0.88 (0.52, 1.48) 0.631 |
| Region | | | |
| Hiran vs. Bay | 1.33 (0.86, 2.06) 0.203 | 1.35 (0.86, 2.11) 0.187 | 1.48 (0.92, 2.40) 0.108 |
| **Intervention** | | | |

| | | | |
|---|---|---|---|
| M2M Groups<br>(Ref: Attended) | 1.27 (0.83, 1.94)<br>0.274 | 1.25 (0.78, 1.99)<br>0.354 | -- |
| NFI Kits<br>(Ref: Received) | *Not assessed due to 81.9% of the sample not receiving NFI kits* | | |
| **Underlying** | | | |
| ***Household Food Security*** | | | |
| Household Hunger Scale<br>(Ref: Little-to-no Hunger) | 1.84 (1.19, 2.82)<br>0.006 | 1.84 (1.13, 2.98)<br>0.014 | 1.79 (1.09, 2.95)<br>0.021 |
| Food Consumption Score<br>(Ref: Acceptable) | 1.68 (1.10, 2.56)<br>0.017 | 1.63 (1.07, 2.50)<br>0.024 | 1.60 (1.04, 2.47)<br>0.032 |
| Reduced Coping Strategy Index<br>(Ref: ≥ 19) | *Not assessed due to 77.5% of the sample having a rCSI <19* | | |
| ***Household Environment*** | | | |
| Household Crowding<br>(Ref: Not Crowded) | 1.21 (0.79, 1.86)<br>0.382 | 1.08 (0.63, 1.83)<br>0.784 | -- |
| Number of CU5<br>(Ref: 1 Child) | 0.82 (0.49, 1.36)<br>0.432 | 0.78 (0.46, 1.30)<br>0.341 | -- |
| Water Treatment<br>(Ref: Treats Water) | *Not assessed due to 78.5% of the sample treating water with an appropriate method* | | |
| Water Source<br>(Ref: Piped) | *Not assessed due to 79.7% of the sample not having household piped water* | | |
| Open Defecation<br>(Ref: Latrine/Toilet) | *Not assessed due to 76.2% of the sample not practicing open defecation* | | |
| Child Stool Disposal<br>(Ref: Latrine/Toilet) | 2.15 (1.28, 3.61)<br>0.004 | 2.23 (1.32, 3.77)<br>0.003 | 2.04 (1.20, 3.49)<br>0.009 |
| Maternal Defecation<br>(Ref: Latrine/Toilet) | 2.23 (1.46, 3.39)<br>0.000 | 2.26 (1.45, 3.52)<br>0.000 | -- |
| ***Health and Nutrition-Related Knowledge and Practices*** | | | |
| Malnutrition Screening<br>(Ref: Mother Screened) | 0.70 (0.44, 1.10)<br>0.124 | 0.74 (0.46, 1.18)<br>0.206 | -- |
| Health-Related Knowledge<br>(Ref: High) | 1.48 (0.96, 2.26)<br>0.073 | 1.44 (0.93, 2.22)<br>0.102 | -- |
| **Immediate** | | | |
| Maternal Age (Ref: 24-34 Years) | | | |
| Younger (<24) | 1.19 (0.71, 1.99)<br>0.520 | 1.21 (0.72, 2.04)<br>0.469 | 1.26 (0.75, 2.14)<br>0.382 |
| Older (35+) | 0.40 (0.23, 0.69)<br>0.001 | 0.39 (0.23, 0.68)<br>0.001 | 0.40 (0.23, 0.70)<br>0.001 |

*Note that final adjusted models may have a smaller sample size due to a few variables missing some observations. **Adjusting for household region and trial arm. ***Adjusting for household region and trial arm. Green indicates statistically significant at p ≤ 0.05. Orange indicates marginally statistically significant from 0.05 < p ≤ 0.10. Pearson goodness-of-fit p-value for final fully adjusted model = 0.6294.

**Table S15.** Endline Overall Maternal MUAC Model: Drivers of Wasting (n=1023)*

| **Domain/Indicator** | **Outcome: Wasting (Mid-upper arm circumference <23 cm of mothers)** | | |
|---|---|---|---|
| | **Unadjusted Bivariate Poisson Regression Coefficient** | **Major Confounder-Adjusted Poisson Regression Coefficient**** | **Final Hierarchical Multivariable Poisson Regression Coefficient***** |
| | Relative Risk Ratio (95% CI) p-value | Relative Risk Ratio (95% CI) p-value | Relative Risk Ratio (95% CI) p-value |
| **Basic/Enabling** | | | |
| ***Wealth and Income Poverty*** | | | |
| Wealth/Asset Index (Ref: Not Poor) | 1.06 (0.74, 1.52) 0.747 | 1.32 (0.80, 2.17) 0.280 | |
| Health- and Nutrition-Related Expenditure (Ref: Middle 50%) | | | |
| Lower 25% | 1.10 (0.72, 1.69) 0.647 | 1.14 (0.74, 1.75) 0.546 | -- |
| Upper 25% | 0.97 (0.62, 1.51) 0.881 | 0.93 (0.59, 1.46) 0.747 | -- |
| ***Household Decision-Making*** | | | |
| Head of Household (Ref: Mother) | 0.99 (0.69, 1.41) 0.948 | 1.01 (0.70, 1.45) 0.975 | -- |
| Decision-Making around Income, Purchases, & Healthcare (Ref: Joint) | | | |
| Maternal | 0.75 (0.47, 1.21) 0.245 | 0.73 (0.45, 1.18) 0.204 | -- |
| Paternal | 0.73 (0.41, 1.30) 0.281 | 0.69 (0.38, 1.25) 0.223 | -- |
| Maternal Education (Ref: Some Education) | 1.25 (0.83, 1.87) 0.283 | 1.25 (0.83, 1.87) 0.289 | -- |
| Displacement (Ref: Not Displaced) | 0.66 (0.36, 1.20) 0.173 | 0.64 (0.34, 1.21) 0.169 | -- |
| Hiran vs. Bay | 1.10 (0.77, 1.57) 0.603 | 1.10 (0.77, 1.58) 0.601 | 1.10 (0.77, 1.58) 0.601 |
| **Intervention** | | | |
| M2M Groups (Ref: Attended) | 0.60 (0.38, 0.94) 0.027 | 0.55 (0.35, 0.87) 0.012 | 0.55 (0.35, 0.87) 0.012 |
| NFI Kits (Ref: Received) | 1.09 (0.75, 1.57) 0.664 | 1.13 (0.77, 1.67) 0.538 | -- |
| **Underlying** | | | |
| ***Household Food Security*** | | | |
| Household Hunger Scale (Ref: Little-to-no Hunger) | 0.97 (0.68, 1.38) 0.855 | 0.94 (0.65, 1.38) 0.764 | -- |
| Food Consumption Score (Ref: Acceptable) | 1.20 (0.84, 1.72) 0.307 | 1.18 (0.82, 1.70) 0.378 | -- |
| Reduced Coping Strategy Index (Ref: <19) | 1.02 (0.69, 1.51) 0.908 | 1.02 (0.69, 1.50) 0.930 | -- |
| ***Household Environment*** | | | |
| Number of CU5 | 1.00 (0.67, 1.51) | 0.97 (0.64, 1.47) | -- |

| | | | |
|---|---|---|---|
| (Ref: 1 Child) | 0.981 | 0.892 | |
| Water Treatment<br>(Ref: Treats Water) | 1.28 (0.80, 2.05)<br>0.304 | 1.30 (0.80, 2.09)<br>0.287 | -- |
| Water Source<br>(Ref: Piped) | 1.12 (0.76, 1.63)<br>0.570 | 1.11 (0.75, 1.64)<br>0.604 | -- |
| Open Defecation<br>(Ref: Latrine/Toilet) | 1.37 (0.94, 2.01)<br>0.103 | 1.36 (0.93, 2.00)<br>0.116 | 1.57 (1.06, 2.33)<br>0.024 |
| Child Stool Disposal<br>(Ref: Latrine/Toilet) | 0.95 (0.66, 1.35)<br>0.758 | 0.95 (0.66, 1.37)<br>0.790 | -- |
| Maternal Defecation<br>(Ref: Latrine/Toilet) | 1.14 (0.80, 1.62)<br>0.482 | 1.10 (0.76, 1.59)<br>0.616 | -- |
| ***Health and Nutrition-Related Knowledge and Practices*** | | | |
| Malnutrition Screening<br>(Ref: Mother Not Screened) | 0.88 (0.60, 1.30)<br>0.531 | 0.98 (0.63, 1.51)<br>0.918 | -- |
| Health-Related Knowledge<br>(Ref: High) | 0.92 (0.63, 1.33)<br>0.657 | 0.85 (0.58, 1.25)<br>0.409 | -- |
| **Immediate** | | | |
| Maternal Age (Ref: 24-34 Years) | | | |
| Younger (<24) | 1.23 (0.77, 1.96)<br>0.382 | 1.23 (0.77, 1.97)<br>0.380 | -- |
| Older (35+) | 0.71 (0.47, 1.07)<br>0.104 | 0.72 (0.48, 1.09)<br>0.120 | -- |

*Note that final adjusted models may have a smaller sample size due to a few variables missing some observations. **Adjusting for household region and trial arm. ***Adjusting for household region and trial arm. Green indicates statistically significant at $p \leq 0.05$. Pearson goodness-of-fit p-value for final fully adjusted model = 0.9975.

45 Venugopal V, Dongre A, Ponnusamy P. Constructing practical and realistic asset-based socioeconomic status assessment scale using principal component analysis for urban

population of Puducherry, India. Indian Journal of Community Medicine. 2021;46:494. doi:10.4103/ijcm.IJCM_925_20.

46 Anderson N, Armeier K, Fuchs S, Khattak Q, Tran A. Multipurpose Outcome Indicators and Guidance. IRC, International Rescue Committee, Mercy Corps, Save the Children International; 2022. Available: https://resourcecentre.savethechildren.net/pdf/Grand-Bargain-MPC-Outcome-Indicators-Guidance.pdf/.

47 Anderson N, Armeier K, Fuchs S, Khattak Q, Tran A. Multipurpose Cash Assistance Monitoring & Evaluation: Baseline - Endline Analysis Guidance Note. IRC, International Rescue Committee, Mercy Corps, Save the Children International. Available: https://resourcecentre.savethechildren.net/pdf/MPCA-Baseline-Endline-Analytical-Guidance-Note.pdf/.

48 Breastfeeding. World Health Organization. Available: https://www.who.int/health-topics/breastfeeding#tab=tab_1. Accessed: 30 September 2024.

49 Infant and young child feeding. World Health Organization. 2020. Available: https://www.who.int/news-room/fact-sheets/detail/infant-and-young-child-feeding#:~:text=WHO%20provides%20simple,%20coherent%20and%20feasible%20guidance%20to%20countries%20for. Accessed: 30 September 2024.

50 Hand Washing Knowledge. People In Need, IndiKit. Available: https://www.indikit.net/indicator/2-wash/67-hand-washing-knowledge. Accessed: 25 September 2-24.

51 Jalata DD, Asefa BG. Trends and Determinants of Dietary Diversity in Children Aged 6–59 Months in Ethiopia: Analysis of 2005–2016 Demographic and Health Survey. Curr Dev Nutr. 2022;6:nzac135. doi:10.1093/cdn/nzac135.

52 Diop L, Becquey E, Turowska Z, Huybregts L, Marie T R, Gelli A. Standard Minimum Dietary Diversity Indicators for Women or Infants and Young Children Are Good Predictors of Adequate Micronutrient Intakes in 24–59-Month-Old Children and Their Nonpregnant Nonbreastfeeding Mothers in Rural Burkina Faso. J Nutr. 2021;151:412–22. doi:10.1093/jn/nxaa360.

53 Castet A. The impact of large-scale land acquisitions on child food insecurity in Africa. World Dev. 2024;179:106597. doi:10.1016/j.worlddev.2024.106597.

54 United Nations Children's Fund. UNICEF Conceptual Framework on Maternal and Child Nutrition. New York, NY, USA: United Nations Children's Fund; 2021. Available: https://www.unicef.org/media/113291/file/UNICEF%20Conceptual%20Framework.pdf.

55 United Nations Children's Fund. Improving child nutrition: The achievable imperative for global progress. New York, NY, USA: United Nations Children's Fund; 2013. Available: https://reliefweb.int/report/world/improving-child-nutrition-achievable-imperative-global-progress.

56 Victora CG, Huttly SR, Fuchs SC, Olinto MT. The role of conceptual frameworks in epidemiological analysis: a hierarchical approach. Int J Epidemiol. 1997;26:224–7. doi:10.1093/ije/26.1.224.

57 Akseer N, Bhatti Z, Mashal T, Soofi S, Moineddin R, Black RE, et al. Geospatial inequalities and determinants of nutritional status among women and children in Afghanistan: an observational study. Lancet Glob Health. 2018;6:e447–59. doi:10.1016/S2214-109X(18)30025-1.

58 Johnston R, Jones K, Manley D. Confounding and collinearity in regression analysis: a cautionary tale and an alternative procedure, illustrated by studies of British voting behaviour. Qual Quant. 2018;52:1957–76. doi:10.1007/s11135-017-0584-6.
